# Understanding the transmission pathways of Lassa Fever: a mathematical modeling approach

**DOI:** 10.1101/2022.09.19.22280113

**Authors:** Praise-God Madueme, Faraimunashe Chirove

**Affiliations:** Department of Mathematics & Applied Mathematics, University of Johannesburg, South Africa

**Keywords:** transmission, dynamics, *Mastomys*, Lassa fever

## Abstract

The spread of Lassa fever infection is increasing in West Africa over the last decade. The impact of this can better be understood when considering the various possible transmission routes. We designed a mathematical model for the epidemiology of Lassa Fever using a system of nonlinear ordinary differential equations to determine the effect of transmission pathways toward the infection progression in humans and rodents including those usually neglected. We analyzed the model and carried out numerical simulations to determine the impact of each of the transmission routes. Our results showed that the burden of Lassa fever infection is increased when all the transmission routes are incorporated and most single transmission routes are less harmful, but when in combination with other transmission routes, they increase the Lassa fever burden. It is therefore important to consider multiple transmission routes to better estimate the Lassa fever burden optimally and in turn determine control strategies targeted at the transmission pathways.

## 1 Introduction

Lassa fever (LF) is an acute viral zoonotic illness responsible for a severe hemorrhagic fever. It is an illness caused by Lassa virus which is a single-stranded RNA virus from the arenavirus family. It was first discovered in 1969 in a Nigerian town called Lassa when two missionary nurses died from the illness. The animal vector of this virus is the multimammate rat (*Mastomys natalensis*) which is one of the most common rodents in West Africa (NICD, 2020; CDC, 2014; Gonzalez, 2020). The *Mastomys* rodents reproduce often and excrete the virus in urine for a very long period of time, and because they occupy human homes, especially where food is stored, they help spread the virus to humans. The transmission of this virus to humans can be through direct or indirect contact. Direct transmissions result from contact between humans and humans, rodents and humans, and rodents and rodents. The evidence available shows that human to human transmission occurs through contact with the body fluids, secretions, excretions, blood of the infectious individual, and sexual transmission (NICD, 2020; Newman, 2018). The infected *Mastomys* rodents are caught (as bush meat) and eaten as food in certain places which directly infects the individuals with the virus. Reports have also shown that there is both horizontal and vertical transmission in multimammate rats especially in seasons when these rodents are actively breeding (Fichet-Calvet et al., 2014; Tewogbola and Aung, Tewogbola and Aung; Gibb et al., 2017). In this paper, however, vertical transmission from rodent to rodent is not covered. Rodents can deposit the virus through their urine and faeces on surfaces in the households such as walls and places where food is stored or even on surfaces where medical equipment is kept. Humans can indirectly acquire the virus when they come in contact with the virus on these contaminated surfaces. Rodents can also become infected because rats share garbage, food on surfaces contaminated with the excretions of infectious rodents yet do not die due to disease but carry the infection and continue to shed it throughout their lifetime (NICD, 2020; Obabiyi and Onifade, 2017). Another form of indirect transmission is through airborne (aerosol) transmission which occurs especially in health centres when people inhale air particles contaminated with the droppings of infected rodents especially during activities like sweeping and other wind activities (NICD, 2020; CDC, 2014; Gonzalez, 2020). The natural history of Lassa fever reveals that its transmission pattern is driven by the frequency of exposure to infected individuals or through contact with infected rodents and contaminated environments (Akhmetzhanov et al., 2019; Sabeti, 2015). It has been shown that Lassa virus is stable as an aerosol, particularly at low relative humidity (30 % RH) and the biological half-life at both 24°C and 32°C ranges from 10.1 to 54.6 minutes (CDC, 2014; Stephenson et al., 1984).

Frequent cases of Lassa virus infection have been seen in endemic regions such as Nigeria, Benin, Ghana, Guinea, Liberia, Mali, Sierra Leone and Togo. Surrounding regions like Central African Republic, Burkina Faso, Côte d’Ivoire, Mali, Senegal, Ghana among others are also at risk, because the rodents that transmit the virus are very common throughout West and East Africa. Hospital staff are also at risk for infection especially in areas with inadequate protective measures and improper sterilization methods (NICD, 2020; Gibb et al., 2017; WHO, 2017). After contracting the virus, humans show symptoms between 1-3 weeks. The presence of virus in the blood is known to peak four to nine days after the onset of symptoms. In most cases, 80% of people infected show no observable or mild symptoms. For these individuals, they show mild signs like slight fever, general malaise and weakness, and headache but do not die due to the infection. Recovery can take place eight to ten days after inception. The remaining 20% of infected individuals can show more severe symptoms like bleeding in the gums, eyes, or nose, respiratory distress, repeated vomiting, facial swelling, pain in the chest, back, and abdomen, shock, and failure in body organs such as liver, spleen and kidneys. The virus in this group of people may also lead to complications such as hearing loss, tremors, encephalitis or even death within 2 weeks after the onset of symptoms due to multiple organ failure (CDC, 2014; WHO, 2017; Yun and Walker, 2012). The number of Lassa virus infections per year in West Africa is estimated at 100,000 to 300,000, with approximately 5,000 deaths. In some places in Liberia and Sierra Leone, the virus led to 10%-16% of people admitted to hospitals every year (NICD, 2020; Gonzalez, 2020; Newman, 2018; Richmond and Baglole, 2003; ACDC, 2018). In 26 out of 36 Nigerian states, a case fatality ratio of 14.8% was recorded between January 1 to February 9, 2020 (WHO, 2020).

Several studies have laid a basis to understand the dynamics of Lassa fever. Some of the studies on Lassa Fever (Onah et al., 2020; Onuorah et al., 2016; Ojo et al., 2021) only considered the basic transmission pathways namely, the human-to-human and the rodent-to-human. Because a great percentage of people show little or no symptoms of Lassa fever, Peter, *et al*. (Peter et al., 2020) described Lassa fever transmission dynamics using a deterministic model integrating the exposed human and rat compartment instead of the usual SIR compartmental structure. Some other studies have tried to establish the time-dependent nature of the transmission dynamics of Lassa fever. For example, Ibrahim and Dénes (Ibrahim and Dénes, 2021) used a compartmental model with time-dependent parameters where the infectious class was partitioned into symptomatically infected, mildly infected and treated individuals alongside with the carrying capacity of the rodent because of the periodic change of weather. Factors like quarantine, hospitalization of infected individuals were also used in (Ibrahim and Dénes, 2021) to comprehend the transmission variability of Lassa fever. In order to understand the epidemiology of the disease, it is important to look at a number of possible transmission pathways through which the virus can be contracted. In this work, we focus on the effects of multiple transmission pathways of Lassa Fever towards the progression of the infection in the human and rodent population. The use of multiple transmission routes gives us a better understanding of the epidemiological structure of Lassa fever. Our study extends the work of Peter *et al* (Peter et al., 2020) and Ibrahim and Dénes (Ibrahim and Dénes, 2021) by:

- Introducing the environment to human transmission pathway. We define the environment as the surfaces, walls and any other equipment where the virus is deposited.
- Introducing the aerosol to human route of transmission. By aerosol, we refer to air particles where the virus is concentrated through human and wind activities.

Thus, our study captures (i) human to human transmission (ii) rodent to human transmission (iii) rodent to rodent transmission (iv) environment to human transmission (v) aerosol to human transmission (vi) environment to rodent transmission. The aforementioned studies form the basic fabric for our work and the understanding gained from them will help us build and analyze a more comprehensive study with more transmission pathways. The remaining part of this work will be arranged thus: Section 2 will be the formulation of the basic model with basic analysis; In Section 3, we will perform our numerical simulations, and discussion and recommendations will be presented in Section 4.

## 2 Model Formulation

The total human population, given as *N*_*H*_ (*t*), is subdivided into five classes which consists of humans susceptible to the virus, *S*_*H*_ (*t*), humans that have Lassa virus but are not infectious, *E*_*H*_ (*t*), infectious humans that are asymptomatic, *I*_*HA*_(*t*), infectious humans that are symptomatic, *I*_*HS*_ (*t*), and humans who have recovered from Lassa fever, *R*_*H*_ (*t*) so that

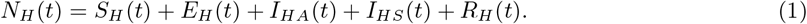

The total rodent population, given as *N*_*R*_(*t*), is subdivided into three classes consisting of rodents susceptible to the virus, *S*_*R*_(*t*), rodents that have Lassa virus but are not infectious, *E*_*R*_(*t*), and infected rodents that can transmit the virus, *I*_*R*_(*t*) with

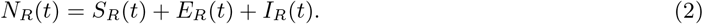

We consider the following direct transmission pathways: the human-to-human contact, the rodent-to-human contact, the rodent-to-rodent contact and the indirect transmission pathway such as the environment-to-human contact and the aerosol-to-human contact and the environment-to-rodent contact. To incorporate the indirect transmission pathways, we use *V*_*S*_ to describe the concentration of Lassa fever virus on the environmental surfaces and *V*_*A*_, the concentration of Lassa virus in the air. The maximum carrying capacity of virus on environmental surfaces and in the air is given by *K*_*V*_, where *V*_*S*_, *V*_*A*_ ≤ *K*_*V*_.

We assume that *π*_1_ is the constant recruitment rate of susceptible humans. The susceptible humans move to the exposed class, *E*_*H*_, through a force of infection

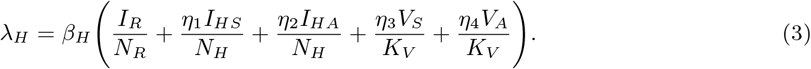

Here, *β*_*H*_ is the effective contact rate between susceptible humans and infected rodents, susceptible humans and infectious humans, susceptible humans, the virus in the environment and the virus in the air, *η*_1_ is the modification parameter which indicates that contact with *I*_*HS*_ is less infectious than with *I*_*R*_. Similarly, *η*_2_, *η*_3_, *η*_4_ are also modification parameters which account for level of infectiousness of contact with *I*_*HA*_, *V*_*S*_ and *V*_*A*_ respectively. Evidence from (Lo Iacono et al., 2015; Lehmann et al., 2017; Bausch et al., 2010; Davies et al., 2019) ensures that the following inequality holds:

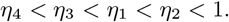

The exposed humans progress to the infectious compartment at the rate *ψ*_1_, where the proportion of exposed individuals that become asymptomatic is *νψ*_1_ and the proportion of exposed persons that become symptomatic is (1 − *ν*)*ψ*_1_. Humans die naturally in all classes at the rate *μ*_1_. Infectious symptomatic humans can die due to the disease at the rate *δ* but there are no cases of death due to infection for the infectious asymptomatic individuals. Infectious asymptomatic and infectious symptomatic humans recover at the rates *ζ*_1_ and *ζ*_2_, respectively.

The susceptible rodents are recruited at a constant rate *π*_2_ and move to the exposed class *E*_*R*_ through a force of infection

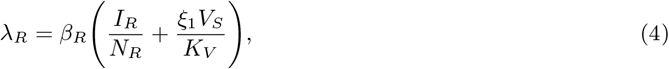

where *β*_*R*_ is the effective contact rate between susceptible rodents and infected rodents and between susceptible rodents and contaminated environment surfaces. *ξ*_1_ is the modification parameter which shows that contact with *V*_*S*_ is less infectious than with *I*_*R*_. The exposed rodents move to the infectious class at the rate *ψ*_2_ and all rodents die naturally at a rate of *μ*_2_. Rodents can also die at a rate *ρ* due to consumption by humans as food. Rodents do not die due to disease since infected rodents can continue to shed the virus throughout their lifetime. The Lassa fever virus is deposited into the environment at rates of *ϕ*_1_, *ϕ*_2_ and *ϕ*_3_ by infectious asymptomatic humans, infectious symptomatic humans, and infected rodents respectively through activities such as urination, excretion of faeces, bleeding and fluid secretions. We further assume that the virus concentration on the environmental surfaces and in the air decays at the rate *θ*_2_ while a proportion of the virus concentration moves into the air at the rate *θ*_3_ through wind and human activities.

The Lassa fever model in Figure 1 is expressed as a system of first order nonlinear ordinary differential equations as follows:

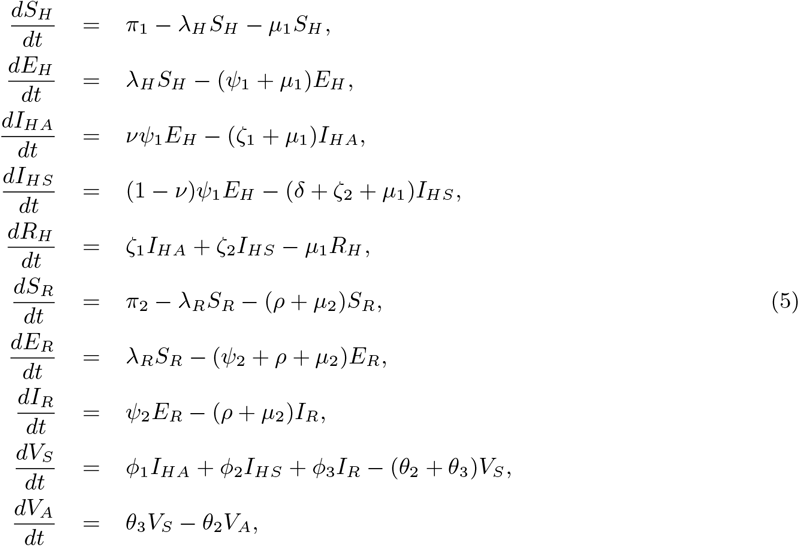

which is subject to the following initial conditions:

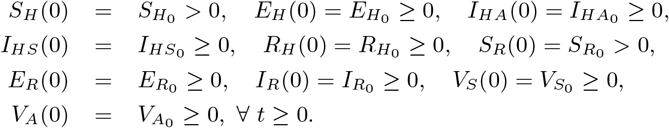

**Figure 1:**
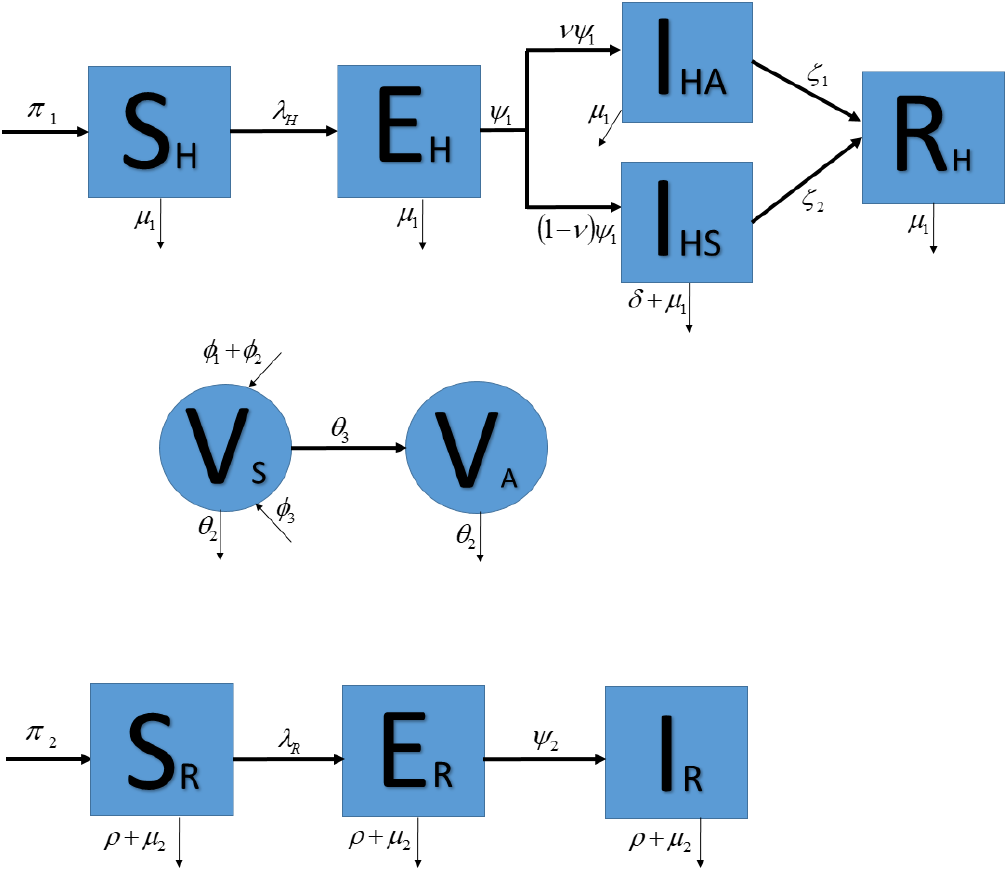
The Lassa fever schematic diagram for human, virus and rodent population.

The variables and parameter descriptions and units are presented in Table 1.

**Table 1:** Description of parameters and variables for model (5)

| Variables | Description | Unit |
| --- | --- | --- |
| $S_H$ | Susceptible human population | people |
| $E_H$ | Exposed human population | people |
| $I_{HA}$ | Infectious asymptomatic human population | people |
| $I_{HS}$ | Infectious symptomatic human population | people |
| $R_H$ | Recovered human population | people |
| $S_R$ | Susceptible rodent population | rodents |
| $E_R$ | Exposed rodent population | rodents |
| $I_R$ | Infected rodent population | rodents |
| $V_S$ | Concentration of Lassa virus in the environmental surfaces | virus |
| $V_A$ | Concentration of Lassa virus in the air | virus |
| Parameters | Description | Unit |
| $\beta_H$ | Contact rate between $S_H$ and $I_R, I_{HA}, I_{HS}, V_S, V_A$ | day<sup>-1</sup> |
| $\eta_1$ | Modification parameter | — |
| $\eta_2$ | Modification parameter | — |
| $\eta_3$ | Modification parameter | — |
| $\eta_4$ | Modification parameter | — |
| $\beta_R$ | Contact rate between $S_R$ and $I_R, V_S$ | day<sup>-1</sup> |
| $\xi_1$ | Modification parameter | — |
| $\psi_1$ | Rate of progression of humans from $E_H$ to $I_{HA}$ and $I_{HS}$ | day<sup>-1</sup> |
| $\psi_2$ | Rate of progression of rodents from $E_R$ to $I_R$ | day<sup>-1</sup> |
| $\theta_2$ | Rate of decay of virus in $V_S$ | day<sup>-1</sup> |
| $\theta_3$ | Rate of progression of virus from $V_S$ to $V_A$ | day<sup>-1</sup> |
| $\phi_1$ | Rate at which virus is shed in $V_S$ by $I_{HA}$ | virus/people × day |
| $\phi_2$ | Rate at which virus is shed in $V_S$ by $I_{HS}$ | virus/people × day |
| $\phi_3$ | Rate at which virus is shed in $V_S$ by $I_R$ | virus/rodents × day |
| $\mu_1$ | Natural death rate of humans | day<sup>-1</sup> |
| $\mu_2$ | Natural death rate of rodents | day<sup>-1</sup> |
| $\rho$ | Death rate of rodents due to consumption by humans | day<sup>-1</sup> |
| $\zeta_1$ | Recovery rate of $I_{HA}$ | day<sup>-1</sup> |
| $\zeta_2$ | Recovery rate of $I_{HS}$ | day<sup>-1</sup> |
| $\pi_1$ | Recruitment rate of humans | people/day |
| $\pi_2$ | Recruitment rate of rodents | rodents/day |
| $K_V$ | Maximum carrying capacity of virus | virus |
| $\delta$ | Disease-induced death rate of humans | day<sup>-1</sup> |
| $\nu$ | Proportion of humans progressing to $I_{HA}$ | — |

### 2.1 Model Analysis

Here, we show that our model is mathematically and biologically meaningful. We shall also compute the basic reproduction number and carry out the stability analysis of the steady states.

#### 2.1.1 Feasible Region

We assume that all parameters are non-negative for time *t* and prove that in the proposed region, Ω, the solutions remain non-negative and bounded. We will analyze the Lassa fever transmission model in the region given as:

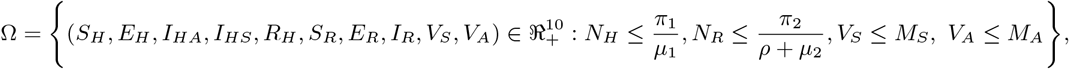

where

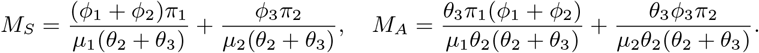

To show that the region Ω is positively invariant, we consider the first equation of (5)

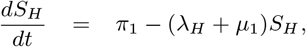

which is solved to obtain

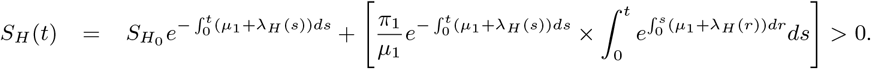

Also, for the solution component of *E*_*H*_ (*t*), we suppose that there exist a first time *t*_1_ such that 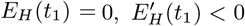 and the rest of the variables are non-negative for 0 *< t*_1_ *< t*. The second equation of system (5) gives

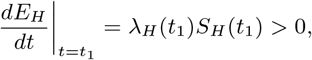

which is a contradiction, so *E*_*H*_ (*t*) ≥ 0, ∀ *t* ≥ 0.

Using a similar approach, it is easy to show that *I*_*HA*_, *I*_*HS*_, *R*_*H*_, *S*_*R*_, *E*_*R*_, *I*_*R*_, *V*_*S*_, *V*_*A*_ are non-negative. Hence, all solutions of (5) are non-negative in Ω.

Now, we show that all solutions with non-negative initial conditions are bounded in the set Ω. It is easy to see that

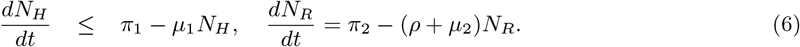

Solving the differential inequality and equation in (6), we use the standard comparison theorem (Lakshmikantham et al., 1989) and the integrating factor to show that as *t* → ∞, we have that 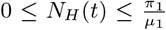 and 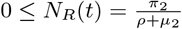. Similarly the differential inequalities

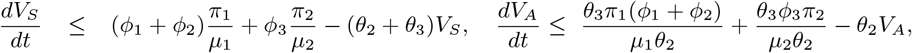

yield 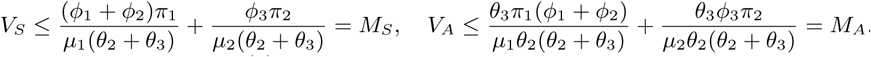

Thus, all possible solutions of (5) enter the region Ω and stay inside it. Hence, the region Ω is positively invariant and attracting and therefore a feasible region.

#### 2.1.2 Reproduction number and equilibria stability analysis

We explore the existence of the equilibrium points of model (5). To obtain the disease free equilibrium (DFE), we equate the right hand side of model (5) to zero and solve when *E*_*H*_ = *I*_*HA*_ = *I*_*HS*_ = *E*_*R*_ = *I*_*R*_ = *V*_*S*_ = *V*_*A*_ = 0 to get:

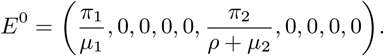

The basic reproduction number *R*_0_ of model (5) is the dominant eigenvalue of the matrix *FV* ^*−*1^ using the next generation matrix approach (Van den Driessche and Watmough, 2002). Here,

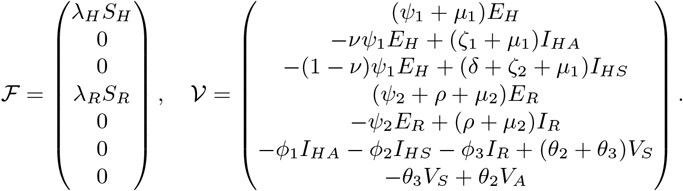

Computing the Jacobian of ℱ and 𝒱 evaluated at *E*^0^ we get

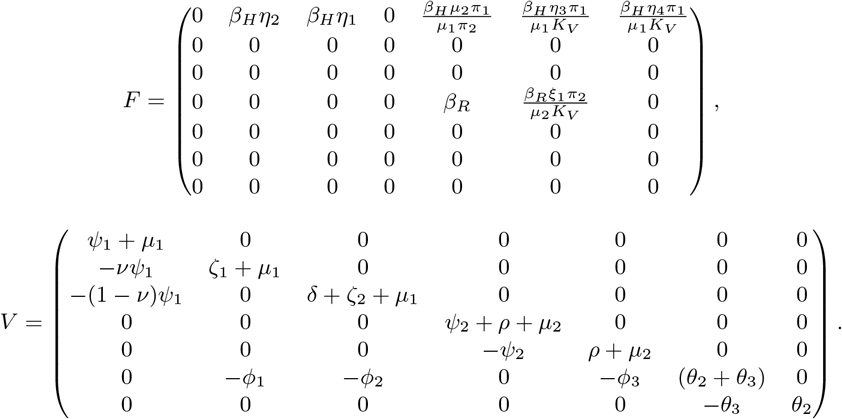

The inverse of *V* is given as

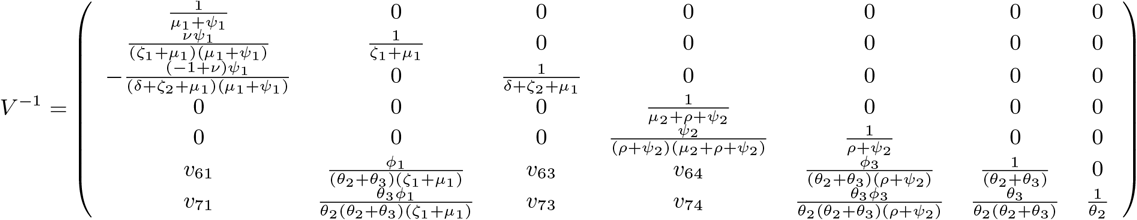

where

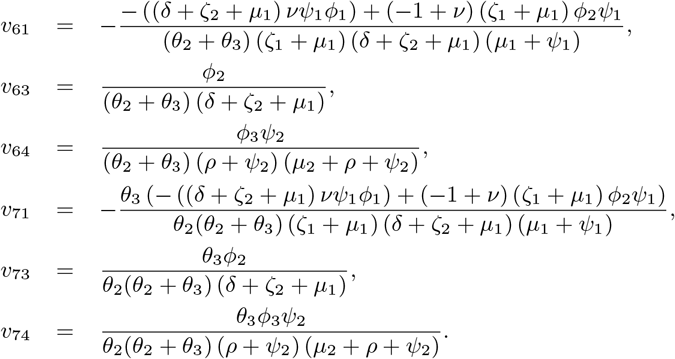

The next generation matrix evaluated at DFE is

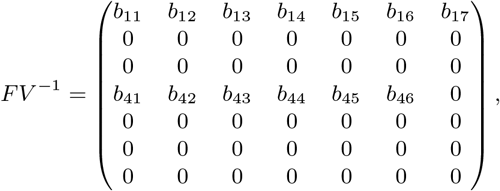

where

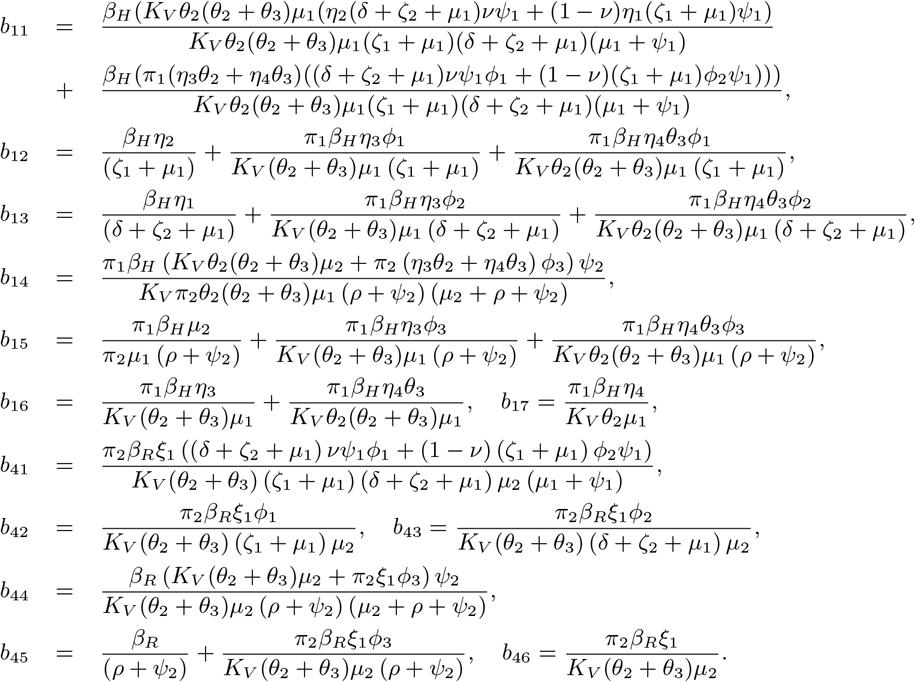

The basic reproduction number is given by

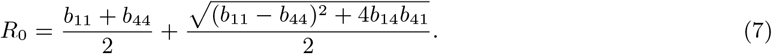

##### Remark 2.1.1.

i. *The terms contained in R*_0_ *represent the direct and indirect transmission pathways. They are described thus: b*_11_ *is the local reproduction number of infectious asymptomatic humans, infectious symptomatic humans, contaminated environmental surfaces and contaminated air particles in the progression of Lassa Fever virus in the human population only; b*_44_ *is the local reproduction number of infected rodents and contaminated environmental surfaces in the progression of Lassa Fever virus in the rodent population only; b*_14_ *is the local reproduction number of contaminated environmental surfaces and contaminated air particles in the progression of Lassa Fever virus in the human population only; and b*_41_ *is the local reproduction number of contaminated environmental surfaces in the progression of Lassa Fever virus in the rodent population only*.
ii. *It is easy to see that*

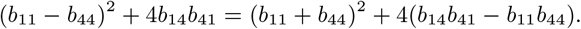 *Thus, b*_14_*b*_41_ *> b*_11_*b*_44_ *implies that R*_0_ *>* 0. *This condition ensures that infection will be sustained across from rodents to humans and from humans to rodents via all the transmission pathways*.
iii. *The local stability of the disease free equilibrium point when R*_0_ *<* 1 *is ensured by the hypothesis used in the computation of R*_0_ *(Van den Driessche and Watmough, 2002)*.

##### Theorem 2.1.1.

*The disease free equilibrium point is globally asymptotically stable if R*_0_ *<* 1.

*Proof*. It suffices to show that our model (5) satisfy conditions **H1** and **H2** of the global stability theorem by (Castillo-Chavez et al., 2002) when *R*_0_ *<* 1.

Model (5) can be rewritten in the form

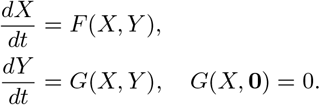

Here, 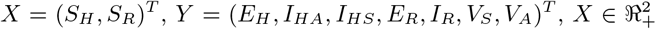represents the number of susceptible humans and rodents and 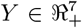 represents the number of exposed humans, asymptomatic infectious humans, symptomatic infectious humans, exposed rodents, infected rodents, contaminated environment and contaminated air.

Our DFE is now written as *E*_0_ = (*X*, **0**) where 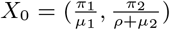 and we show that the following conditions are satisfied:

- **H1**: For 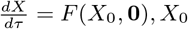, *X*_0_ is globally asymptotically stable.
- **H2**: *G*(*X, Y*) = *AY* − *Ĝ*(*X, Y*), *Ĝ*(*X, Y*) ≥ 0 for (*X, Y*) ∈ Ω where *A* = *D*_*Y*_ (*G*(*X*_0_, **0**)) is an M-matrix and Ω is the region where the model makes biological sense.

For the first condition **H1**, we have

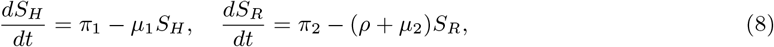

which can be solved to get

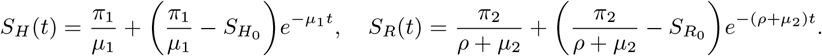

We take limits as *t* → ∞ to get,

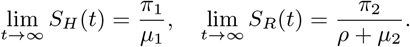

Hence, the solutions of equation (8) converge to *X*_0_ regardless of the initial conditions. Therefore, *X*_0_ is a globally asymptotically equilibrium point of (8).

For condition **H2**, we consider *F* (*X*, 0) = (*π*_1_ − *μ*_1_*S*_*H*_, *π*_2_ − (*ρ* + *μ*_2_)*S*_*R*_), *G*(*X, Y*) = *AY* − *Ĝ*(*X, Y*) where

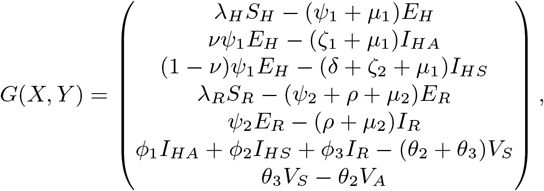

and

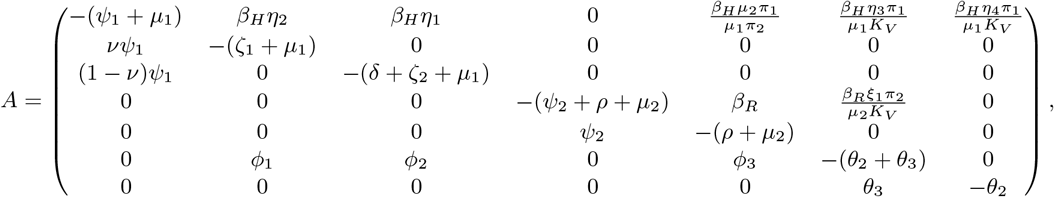

which is an M-matrix and

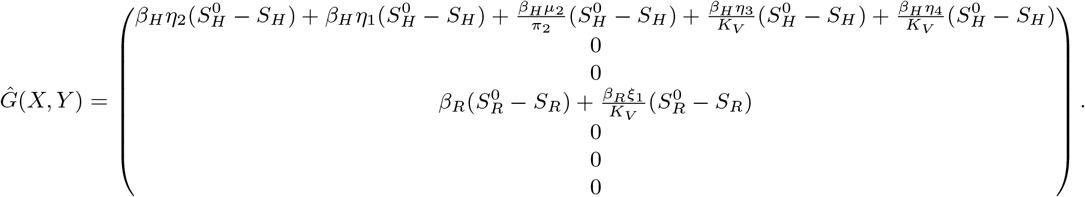

Clearly, *Ĝ*(*X, Y*) ≥ 0 since 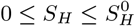 and 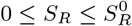. □

The global stability of *E*^0^ thus follows and the Lassa fever virus can be eliminated from the human and rodent population over a period of time provided *R*_0_ *<* 1.

The endemic equilibrium point of the model (5) is given by

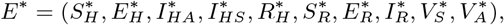

where

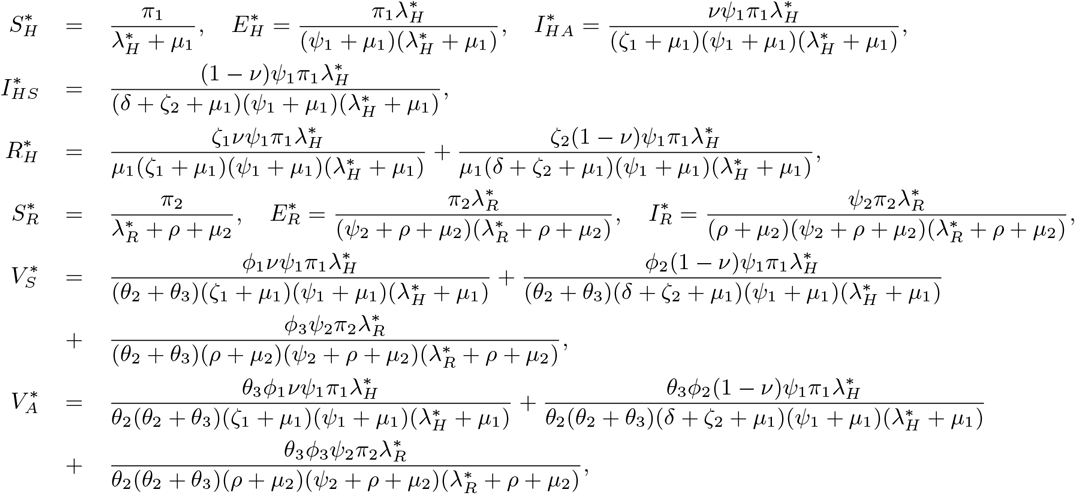

and

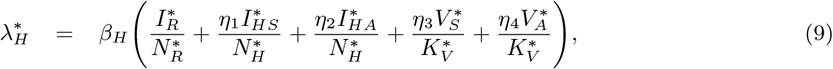

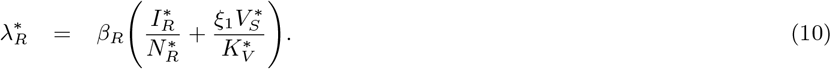

Equations (9) and (10) can be written explicitly as

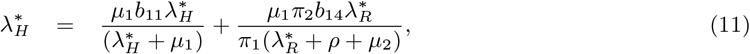

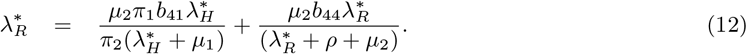

We see that the state variables are expressed in terms of 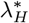and 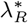. From here, we proceed by using the approach in (Moghadas et al., 2003; Velasco-Hernandez and Hsieh, 1994). Hence, we can obtain positive equilibrium points of the model by finding the fixed points of equations (11) and (12) as

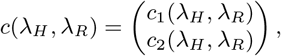

where

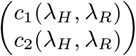

corresponds to the right hand sides of equation (11) and (12).

##### Theorem 2.1.2.

*There exists a unique fixed point* 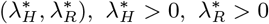 *which satisfies*

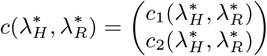

*and corresponds to the endemic equilibrium point E*^*\**^ *(Moghadas et al*., *2003; Velasco-Hernandez and Hsieh, 1994)*.

*Proof*. From the first equation, we fix *λ*_*R*_ *>* 0 and look at the real-valued function depending on *λ*_*H*_ :

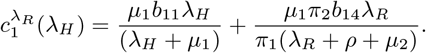

We have that

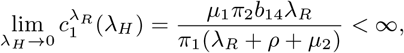

and

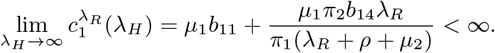

It follows that 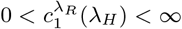 which implies that 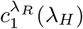 is bounded for every fixed *λ*_*R*_ *>* 0. Next,

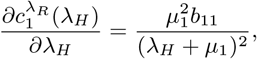

and

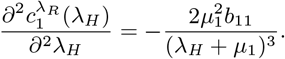

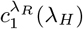 is an increasing concave down function since 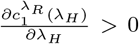 and 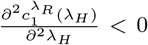. Hence, there is no change in concavity of *c*_1_ in the bounded domain. It follows that there exists a unique 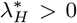 which satisfies 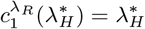.

For this 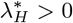, we look at the real-valued function depending on *λ*_*R*_:

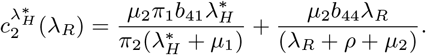

Then,

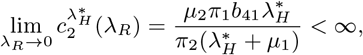

and

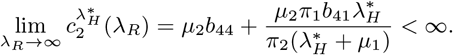

It follows that 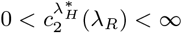 which implies that 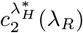 is bounded for every fixed 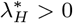.

Next,

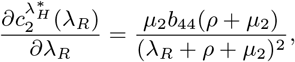

and

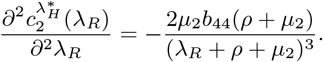

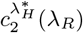 is an increasing concave down function since 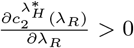 and 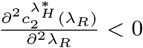. Hence, there is no change in the concavity of *c*_2_ in the positive domain. It follows that there exists a unique 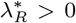 which satisfies 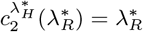.

Therefore, there is a fixed point 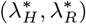 which corresponds to the endemic equilibrium point *E*^*\**^. □

We now investigate the stability of the equilibrium points using the stability of the fixed point system 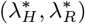 corresponding to *E*^*\**^. The Jacobian of the system is given by:

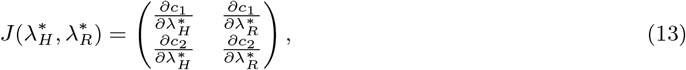

where

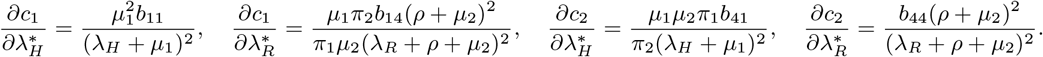

We note that 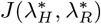 evaluated at the fixed point, 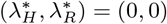, is given by

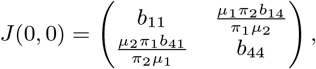

and for stability, we require that |*λ*_*i*_| *<* 1, where *λ*_*i*_ are the eigenvalues of *J* (0, 0), which corresponds to

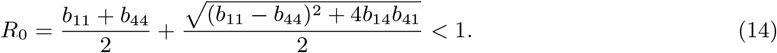

Hence, the stability of 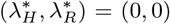 is achieved when *R*_0_ *<* 1. The point is unstable provided *R*_0_ *>* 1. Thus, the stability of 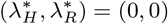 corresponds to the stability of *E*^0^ when *R*_0_ *<* 1. Now, for 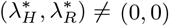, we have

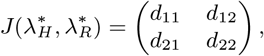

and for stability, we require that |*λ*_*i*_| *<* 1, that is,

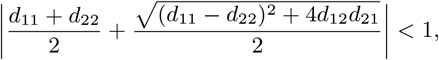

where

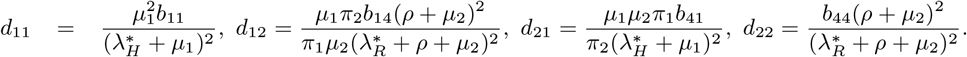

The stability of the fixed point system is governed by the fact that the absolute value of the eigenvalues of the fixed point system is less than unity (Velasco-Hernandez and Hsieh, 1994; Moghadas et al., 2003). Hence, |*λ*_*i*_| *<* 1 corresponds to

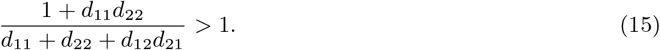

Defining the left hand side of (15) as 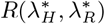, the fixed point 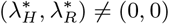 is stable when 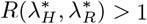.

## 3 Numerical Simulations

### 3.1 Parameter estimation

It is crucial to estimate the model parameter values in order for us to perform numerical analysis. We consider the ecological niche where Lassa fever is endemic. We focus on three (3) states in Nigeria (Ondo, Edo and Ebonyi) where this virus has ravaged communities in the past few years based on Nigeria Centre for Disease Control (NCDC) reports (NCDC, 2021).

In the chosen region, we consider a few local sites in the three states of about 10000 persons, 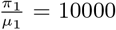. The human natural death rate is 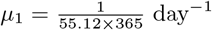 using the average human lifespan in Nigeria as 55.12 (Trends, 2021). The daily recruitment rate of humans is estimated as *π*_1_ = 10000 × *μ*_1_ = 0.497 day^*−*1^. The value, *ν* = 0.8 since 80% of individuals are asymptomatic. The sample study of Lassa fever cases in Nigeria shows a case fatality ratio of 18.9% in the year 2021 (NCDC, 2021), we assume *δ* = 0.189 year^*−*1^ which translates to *δ* = 0.0005 day^*−*1^. Research conducted in these communities had reported a yearly rodent consumption rate of 29.9% in Edo State, 11% in Ebonyi State and 20.2% in Ondo State (WHO, 2021; Ossai et al., 2020). So, we use an average consumption rate of 20.4% per year giving us *ρ* = 0.0006 day^*−*1^. The biological half-life of Lassa virus ranges from 10.1 to 54.6 minutes (Stephenson et al., 1984); so using 10.1 minutes implies that 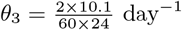.

We consider a hypothetical average population of *Mastomys* rat to be 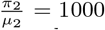, since there is no known quantified estimate of the rodents population. The average lifespan of a rodent is 1 year (Control, 2018), so we obtain 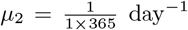. We estimate the rodent recruitment rate to be *π*_2_ = 1000 × *μ*_2_ = 2.74 day^*−*1^. Some of the parameters cannot be found or estimated from literature, so we used model calibration to get ideal representation curves for all state variables to get approximate values. Thus *ψ*_1_ = 0.0094 day^*−*1^, *ψ*_2_ = 0.048 day^*−*1^, *β*_*H*_ = 0.00017 day^*−*1^, *β*_*R*_ = 0.004 day^*−*1^, *ξ* = 0.167, 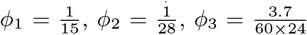 and the values of *η*_1_, *η*_2_, *η*_3_, *η*_4_, to lie in the interval (0, 1). For our simulation, we use the following initial conditions: *S*_*H*_ (0) = 10000, *E*_*H*_ (0) = 0, *I*_*HA*_(0) = 324, *I*_*HS*_ (0) = 81, *R*_*H*_ (0) = 10, *S*_*R*_(0) = 1000, *E*_*R*_(0) = 0, *I*_*R*_(0) = 100, *V*_*S*_ (0) = 1000, *V*_*A*_(0) = 100. Table 2 contains the parameter values used in the simulations.

**Table 2:**
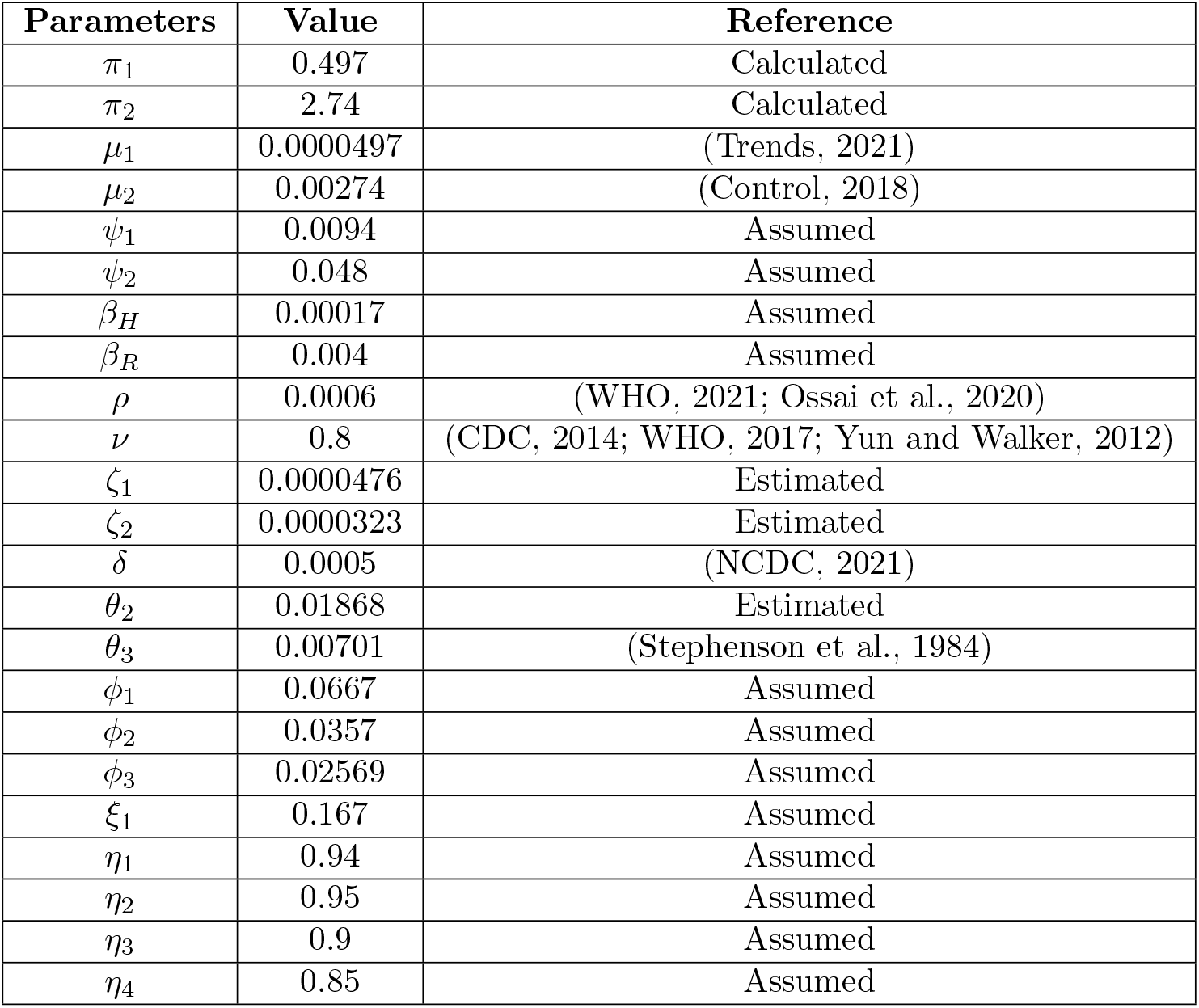
Parameter values and references

### 3.2 Sensitivity Analysis

Sensitivity analysis is a procedure used to determine the strength of model predictions to parameter values. It is crucial because there are usually flaws in assumed parameter values and generally in data collection. Sensitivity analysis shows the parameters that deserve the best numerical attention, reveals insensitive parameters that do not require much effort to estimate and shows which parameters should be targeted for intervention (Mikucki, 2012). Local sensitivity analysis is based on calculating the effect on the model output of small perturbations around a nominal parameter value. This perturbation is done on one parameter at a time using the first-order partial derivative of the model output with respect to the perturbed parameter. Here, we will investigate parameters that have a high impact on *R*_0_, and should be targeted by intervention strategies. The global sensitivity analysis, on the other hand, seeks to explore the input parameters space across its range of variation and then quantify the input parameter importance based on a characterization of the resulting output response surface. It is a sampling-based method that investigates uncertainties for parameter values in the entire parameter range (Saltelli et al., 2008, 2004; Marino et al., 2008; Turányi, 1990; Blower and Dowlatabadi, 1994). We will perform both the local and global sensitivity analysis.

#### 3.2.1 Local Sensitivity Analysis of *R*_0_

We calculate the local sensitivity indices of the parameters with respect to *R*_0_ using the normalized forward index. These indices reveal the importance of each parameter to disease transmission and should be taken into consideration while defining our control strategies. According to (Chitnis et al., 2008), the normalized forward sensitivity index of a variable *u* that depends differentiably on a parameter *ρ* is defined as:

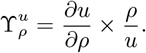

For example, the sensitivity index of *R*_0_ with respect to *β*_*H*_ will be

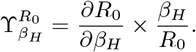

When 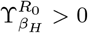, we say that *β*_*H*_ increases the value of *R*_0_ as its value increases, while if 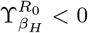, then *β*_*H*_ decreases the value of *R*_0_ as its value increases. The results of the sensitivity indices is shown in table 3.

**Table 3:** Sensitivity indices of *R*_0_.

| Parameters | Sensitivity indices of $R_0$ | Parameters | Sensitivity indices of $R_0$ |
| --- | --- | --- | --- |
| $\pi_1$ | 0.444782 | $\delta$ | -0.0272335 |
| $\pi_2$ | 0.0000581831 | $\theta_2$ | -0.439338 |
| $\mu_1$ | -0.946283 | $\theta_3$ | -0.00550217 |
| $\mu_2$ | -0.000168322 | $\phi_1$ | 0.43505 |
| $\psi_1$ | 0.00524857 | $\phi_2$ | 0.00973221 |
| $\psi_2$ | -0.00190397 | $\phi_3$ | 0.0000581831 |
| $\beta_H$ | 0.997936 | $\xi_1$ | 0.00199971 |
| $\beta_R$ | 0.0020637 | $\eta_1$ | 0.0219675 |
| $\rho$ | -0.0000495958 | $\eta_2$ | 0.531187 |
| $\nu$ | 0.839438 | $\eta_3$ | 0.32696 |
| $\zeta_1$ | -0.472691 | $\eta_4$ | 0.115881 |
| $\zeta_2$ | -0.00175928 | | |

We observe from table 3 that parameters such as *β*_*H*_, *ν, η*_2_, *π*_1_, *ϕ*_1_, *η*_3_, *η*_4_, *η*_1_, *β*_*R*_, *ξ*_1_ are positively correlated with *R*_0_ thus increase in these parameters increase the reproduction number. The parameters *μ*_1_, *ζ*_1_, *δ, θ*_2_ are negatively correlated with *R*_0_ thus they decrease the value of *R*_0_ as they are increased. There are parameters such as *π*_2_, *μ*_2_, *ρ, ϕ*_3_ that are insensitive with respect to the reproduction number of Lassa fever in the population. These parameters do not require too much effort to estimate and will not cause much changes to *R*_0_ when they are increased or decreased. All parameters associated with infection pathways have positive indices and thus, all the infection pathways have a potential to collectively or otherwise increase the infection. We also observe that *β*_*H*_ is the most sensitive parameter followed by *ν, η*_2_, *π*_1_, *ϕ*_1_, *η*_3_, *η*_4_, *η*_1_, *β*_*R*_, *ξ*_1_ respectively. Intervention strategies can be targeted at reducing the impact of parameters which increase *R*_0_ whilst increasing those that reduce it.

#### 3.2.2 Global Sensitivity Analysis

The global sensitivity analysis is carried out using the Latin Hypercube Sampling and Partial Rank Correlation Coefficients (PRCCs) (Marino et al., 2008). This is a robust sensitivity measure that combines uncertainty analysis with partial correlation on rank-transformed data to assess the sensitivity of our outcome variable to parameter variation. Figure 2 shows that the parameters *β*_*H*_, *β*_*R*_, *ϕ*_1_, *ϕ*_3_, *θ*_3_, *ψ*_1_, *ν* are positively correlated to *C*_*H*_ and thus increase the burden of Lassa fever infection in the human population; the parameters *θ*_2_, *μ*_2_, *π*_1_ are negatively correlated to *C*_*H*_ and decrease the burden of Lassa virus in the human population when they are increased. There are also insensitive parameters *ψ*_2_, *μ*_1_, *ψ*_1_, *ζ*_1_, *ζ*_2_, *ϕ*_2_ with PRCCs very close to zero. The parameters associated with infection pathways though weakly correlated, remain positively correlated in the different populations. In the rodent population, the parameters *β*_*H*_, *β*_*R*_, *ψ*_1_, *ψ*_2_, *μ*_1_, *μ*_2_, *θ*_3_ are positively correlated to *C*_*R*_ and increase the infection burden in rodents, parameters *π*_1_, *π*_2_, *μ*_2_, *ϕ*_2_, *ρ, μ*_2_, *θ*_3_ are negatively correlated to *C*_*R*_ while there are parameters with PRCCs close to zero such as *δ, θ*_3_, *ζ*_1_, *ζ*_2_ and so on. From the PRCCs of the virus population, we see positively correlated parameters such as *β*_*R*_, *ψ*_2_, *π*_2_, *μ*_1_, *ξ*_1_, negatively correlated parameters *μ*_2_, *ρ, π*_1_ and other parameters with PRCCs very close to zero. In all populations, we see that some parameters are positively or negatively correlated at certain time points but become insensitive at other time points and verse visa. Hence, the interplay and the exchange of sensitivity by different parameters on different variables alludes to the complexities brought about by the multiple transmission pathways which in turn suggest the importance of every pathway in the prognosis of Lassa fever.

**Figure 2:**
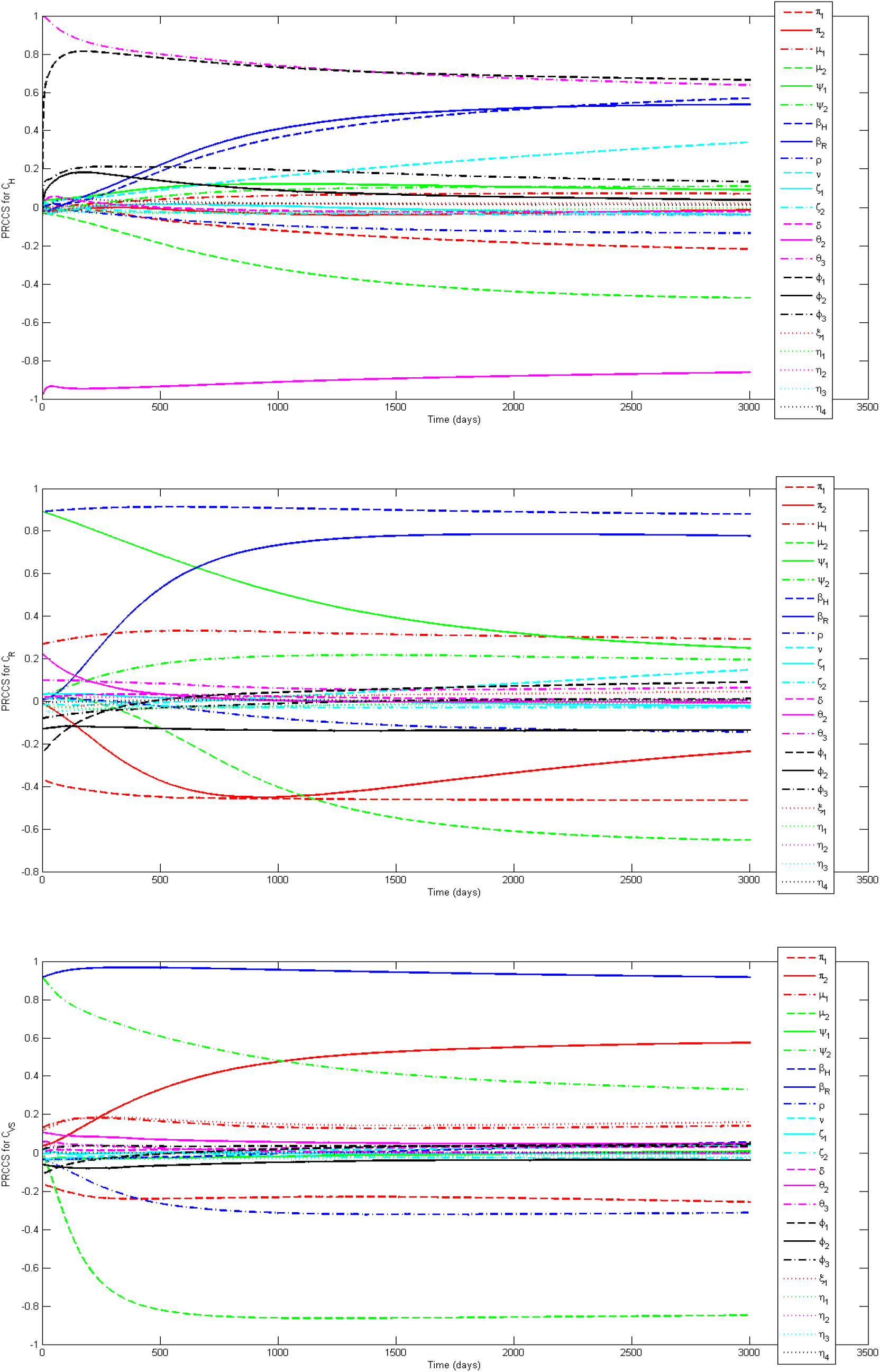
Partial Rank Correlation Coefficient for the full range of parameters of model (5).

### 3.3 Simulation results

Figure 3 shows the baseline graphs of the system (5) without varying the system parameters. The simulations were done over a time period of 40000 days. The baseline graph is perceived to represent the ideal situation where Lassa fever persists in the system. We will illustrate the impact of the transmission pathways in the next subsection.

**Figure 3:**
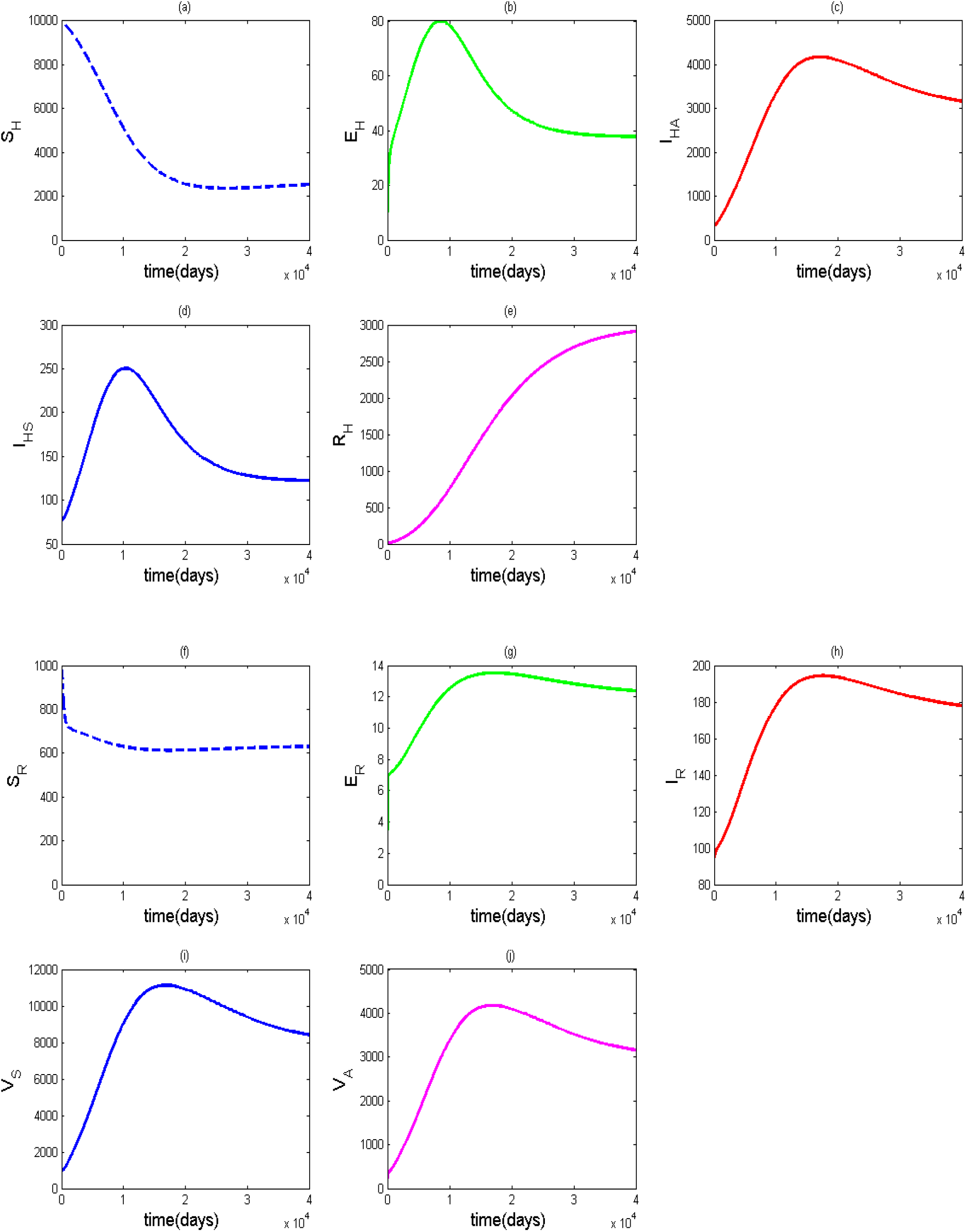
Graphical illustration of model (5) for *R*_0_ greater than unity: *R*_0_ = 3.1301.

#### 3.3.1 Simulation of the transmission pathways

We now investigate the impact of the various transmission routes on the progression of Lassa fever in both human and rodent population as well as the growth of virus in the environment. We shall proceed using the following strategies:

1. Transmission pathways for the human population
  a. 5 Single transmission pathways (see Figure 4).

**Figure 4:**
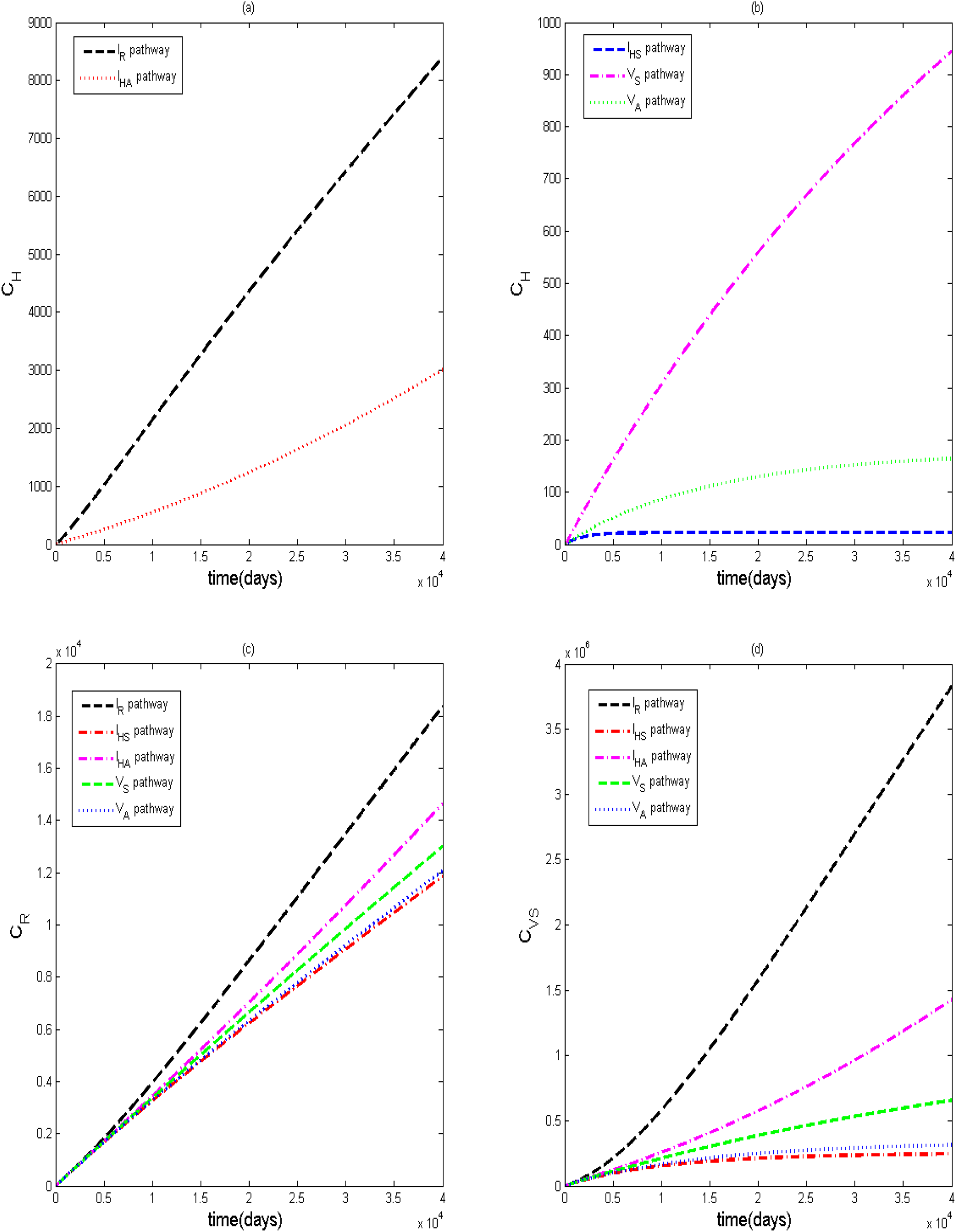
Graphical illustration of model (5) for single transmission routes
  b. 10 combinations of two transmission pathways (see Figure 5).

**Figure 5:**
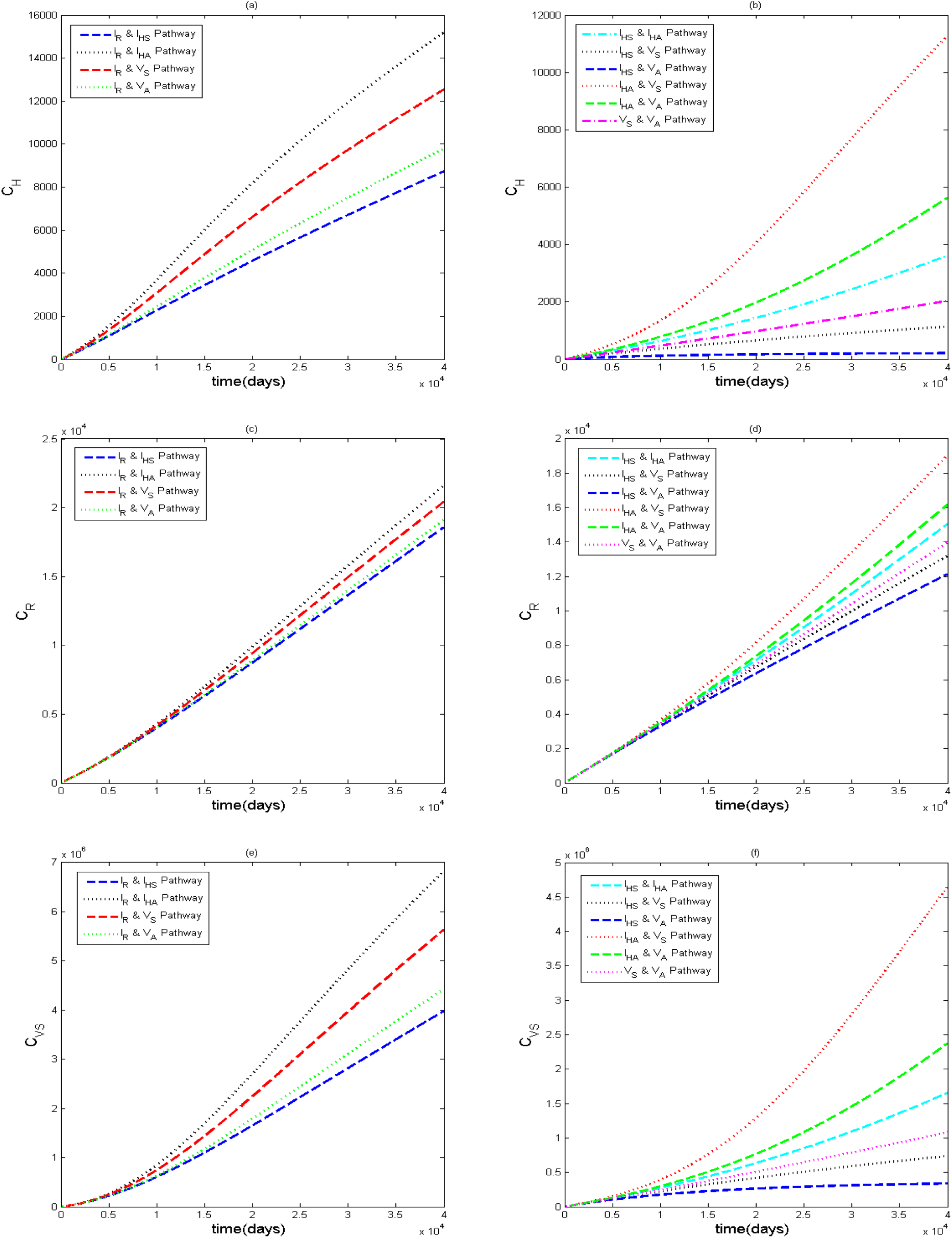
Graphical illustration of model (5) for double transmission routes on human, rodent and virus classes
  c. 10 combinations of three transmission pathways (see Figure 6).

**Figure 6:**
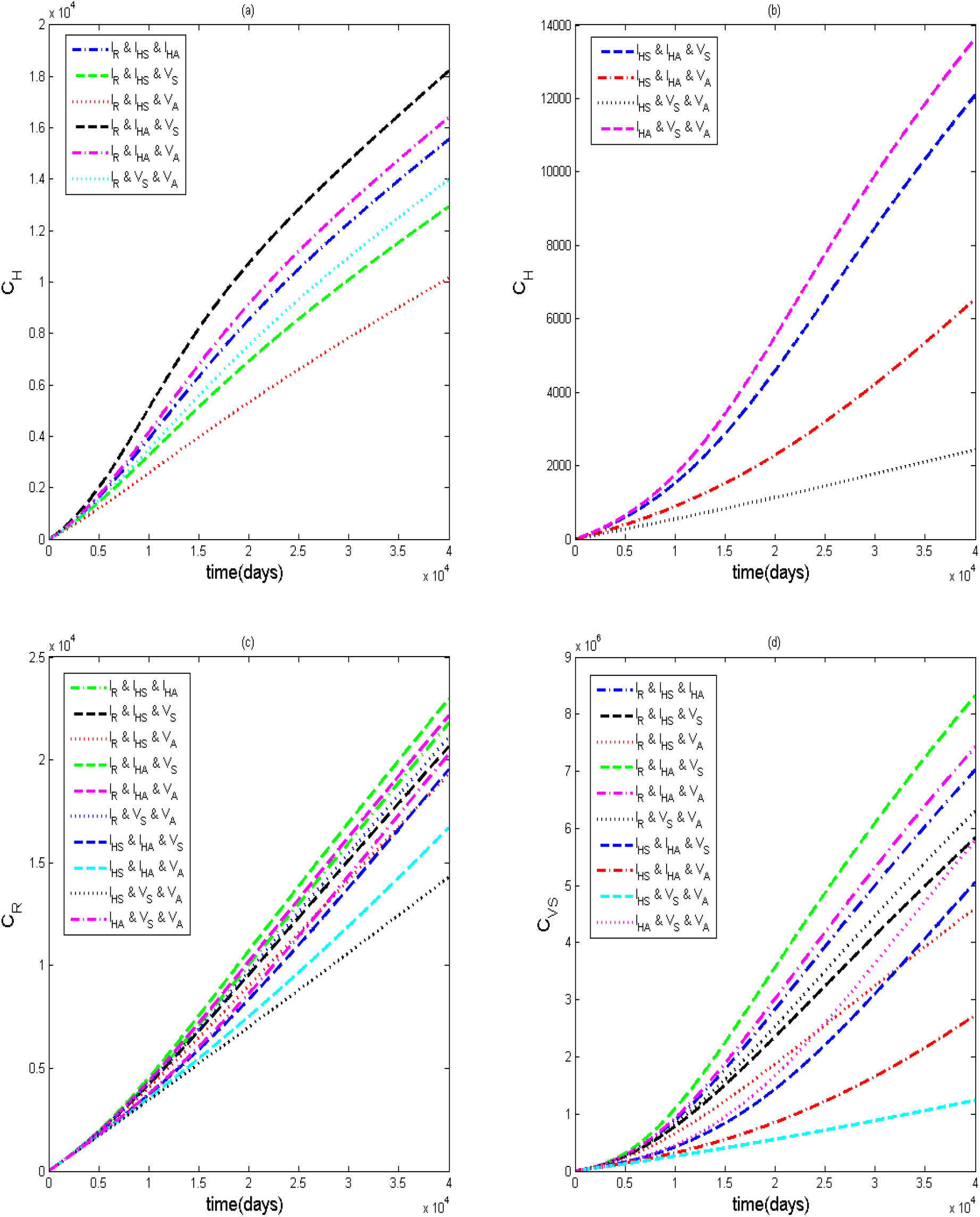
Graphical illustration of model (5) for three transmission routes on human, rodent and virus classes
  d. 5 combinations of four transmission pathways (see Figure 7).

**Figure 7:**
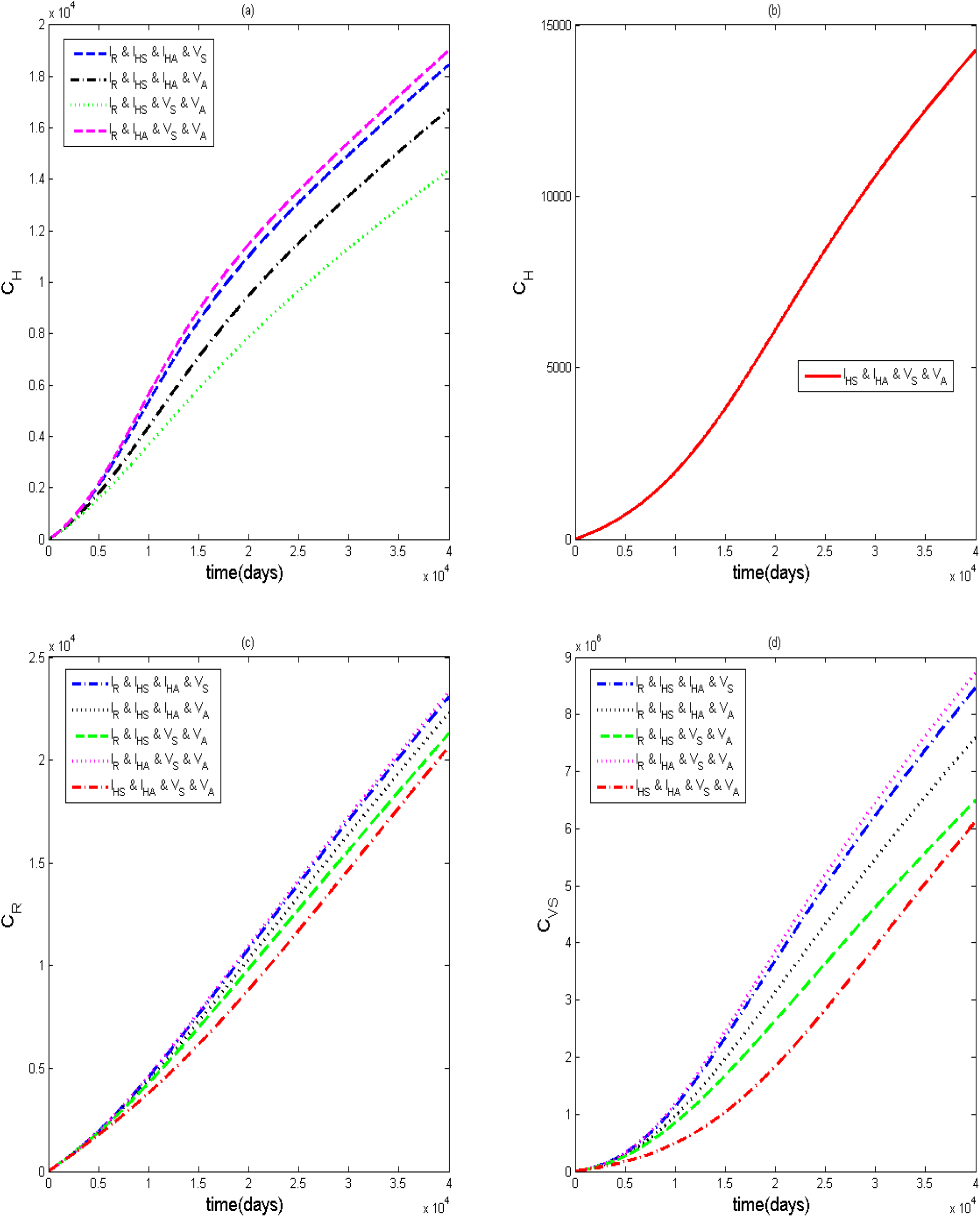
Graphical illustration of model (5) for four transmission routes on human, rodent and virus classes
  e. 1 combination of five transmission pathways (see Figure 8).

**Figure 8:**
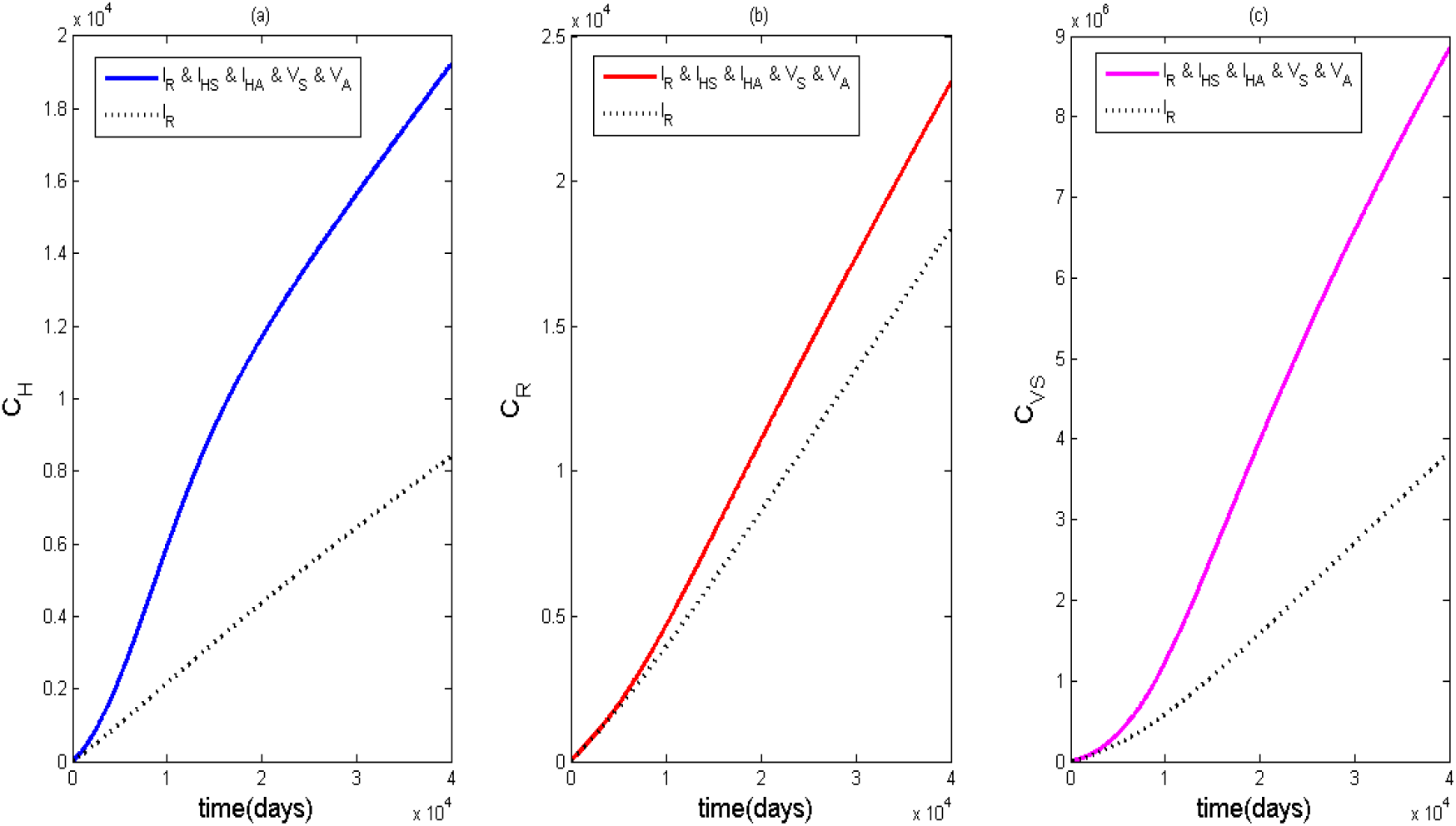
Graphical illustration of model (5) for all transmission routes
2. Transmission pathways for rodent population
  a. Single transmission pathways (see Figure 8).
  b. 1 combination of two transmission pathways (see Figure 9).

**Figure 9:**
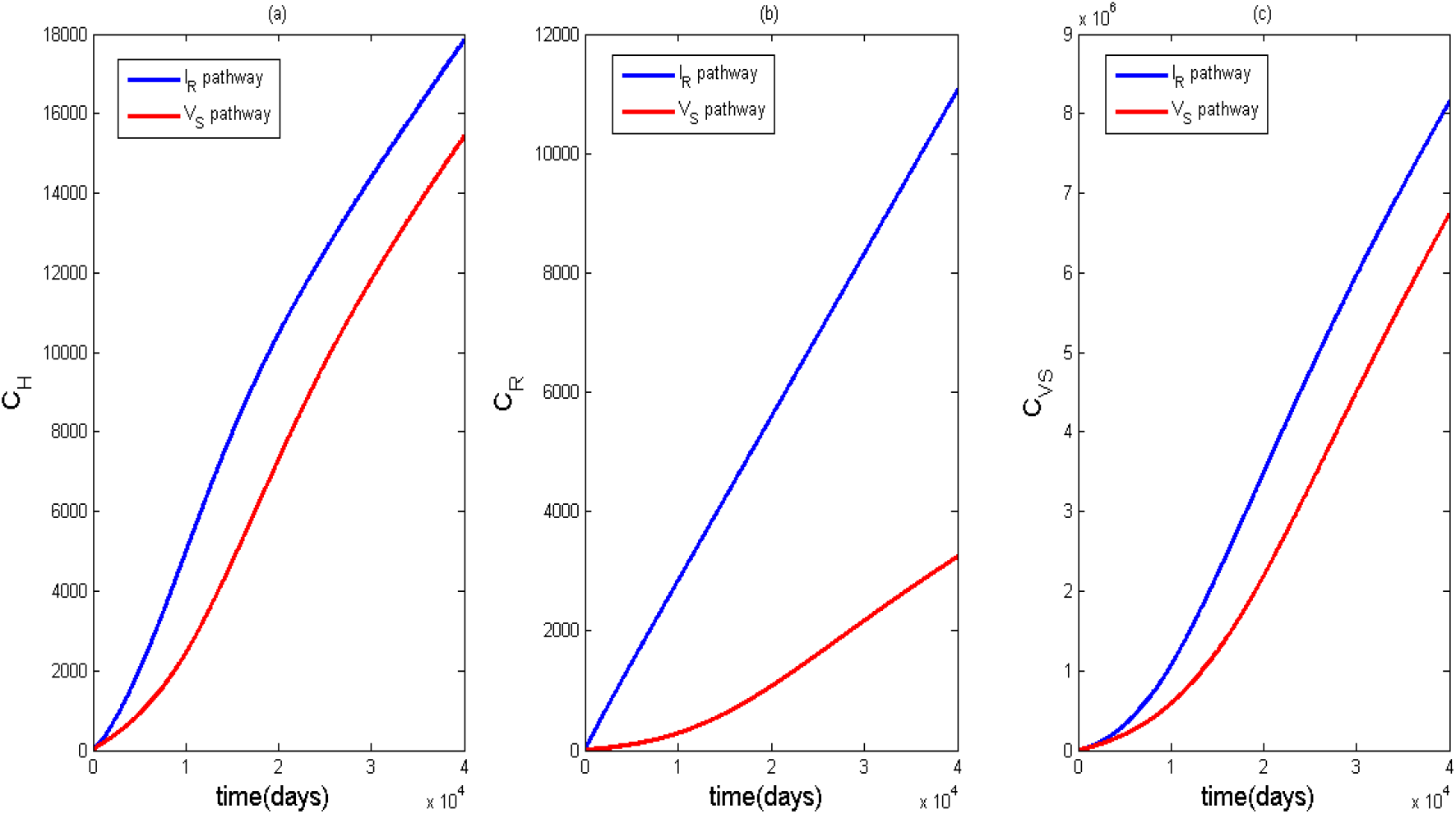
Graphical illustration of model (5) for rodent transmission routes on all the classes.

By single transmission pathway, we use each of the single contact rates in the human force of infection and test its impact on the system while the entire rodent force of infection is operational. We do the same for two transmission routes and continue till we exhaust all other transmission pathways. We also investigate using each of the single contact rates in the rodent force of infection and the two transmission routes while keeping the entire human force of infection in use. We shall test these strategies using the cumulative cases (NCDC, 2021) in the humans, rodents and virus in the environment. To capture this, we will simulate the cumulative cases using the equations:

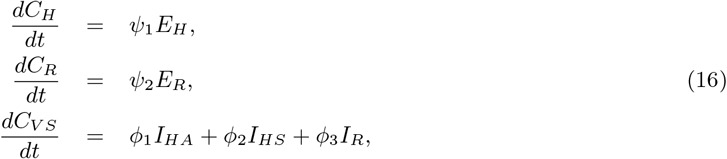

subject to the initial conditions *C*_*H*_ (0) = 0, *C*_*R*_(0) = 0, 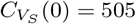, where *C*_*H*_ is the cumulative infection cases in the human population, *C*_*R*_ is the cumulative infection cases in the rodent population, and *C*_*V S*_ is the cumulative infection cases in the virus population.

Figure 4 reveals that the effective contact rate between susceptible humans and infected rodents does the most damage with regards to the progression of infection. This is followed by the contact rate between susceptible humans and infectious asymptomatic humans which is less infectious and then by the contact with contaminated environment, contaminated air, and infectious symptomatic humans. We observe that every single route of transmission plays a role in driving the Lassa fever infection even though some are less significant than the others. In the rodent population, we also see a notable difference in the level of infectiousness of the transmission routes likewise in the virus population. This shows that some pathways are more deadly than others yet every pathway makes their own contribution. We see from Figure 5 that a combination of two transmission pathways increases disease burden more than a single pathway. We also see that some combinations are more deadly than others. Any combination with the effective contact rate between susceptible humans and infected rodents produces a surge of infections followed by any combination with the effective contact rate between susceptible humans and infectious asymptomatic humans and then other pathways. Overall, we see that as the number of transmission routes increase, the burden of infection increases also (see figures 6-7). Figure 8 shows a combination of all the transmission routes plotted alongside the dominant single transmission pathway. The region between the two graphs accounts for the contribution of other pathways in combination with the effective contact rate between susceptible humans and infected rodents. This shows that even though the effective contact rate between susceptible humans and infected rodents is dominant, other pathways should not be neglected because when they work in combination, there is an additional increase in the burden of Lassa fever over a cumulative period of time. It is also important to note that horizontal transmissions between susceptible rodents and infected rodents also play a huge role in increasing the infection as well as contact rate between susceptible rodent and contaminated environmental surfaces (see figure 9). We use a table to illustrate the effect of the multiple transmission routes (see table 4) in appendix A. The values in the table are collected at the end point of simulations.

### 3.4 Discussion of Results

We investigated the transmission dynamics of Lassa fever infection incorporating multiple transmission routes to capture their impact on the progression of the infection. Using a deterministic model that accounts for Lassa fever infection, we were able to show how incorporating several transmission pathways affects the prevalence of the disease. We used some mathematical tools to establish the local stability of the endemic equilibrium and the global stability of the disease free equilibrium. From our analysis, we got mathematical expressions that shows the conditions for which the disease will persist or be controlled in the system and illustrated sensitivity of parameters changes as system dynamics progress.

From our model simulations, we see that every transmission pathway has an impact towards the progression of Lassa fever. However, there are some routes of transmission that contribute significantly more than others. Control measures should be targeted more on the contact rates between susceptible humans and infected rodents (especially in areas where rodent consumption is high), and contact rates between susceptible humans and infectious asymptomatic humans. A great challenge arises when dealing with susceptible and asymptomatic infected humans pathway because they are not easily identified through symptoms. This calls for control methods that can detect this category of people such as mass testings in endemic areas, vaccination and so on. It is also important not to neglect the contact rates between susceptible humans and contaminated air particles especially in health centres with recorded Lassa fever cases and the contact rates between susceptible humans and contaminated environment (especially in poorly sanitized areas) because they are further drivers of infection (CDC, 2014).

Most single transmission routes are less harmful, but when they operate in combination with other transmission routes, they contribute additional damage to the system. Studies (Onah et al., 2020; Ojo et al., 2021; Peter et al., 2020; Ibrahim and Dénes, 2021) that only concentrated on the human and rodent direct transmission routes have not captured valuable information on the environmental impact towards the progression of the infection. This work gives a more comprehensive breakdown of the transmission dynamics of Lassa fever as it integrates indirect transmission routes which are sometimes neglected but play a crucial role in increasing the infection statistics. Current reports show that about four medical doctors died and 38 health workers were infected during a recent Lassa fever outbreak in Nigeria (Punch, 2022; NCDC, 2022). This increase in the death of health workers tells of the fact that serious measures should be taken to curb the spread of the virus through the indirect transmission routes like the environmental surfaces and aerosol. This will help to reduce the impact of these indirect transmission routes on the infection chain. Further studies can be targeted at (i) combination of multiple routes of transmission incorporating the effect of seasonality of infection, (ii) proper sanitation, intervention strategies and holistic control measures that integrate these multiple transmission pathways which can help public health reduce disease prevalence, (iii) optimizing cost of several control measures using Cost Effective Analysis so that individuals in endemic areas with issues of poverty can be properly assisted. Access to real field data can also improve the predictive capacity of the current model. Alternative techniques like the scaling of the model can also be used to help simplify the analysis where parameters are dimensionless and express ratios of physical effects rather than levels of individual effects(Ledder, 2017).

## Data Availability

All data produced in this manuscript are contained in the manuscript

## A Appendix Section

**Table 4:** The outputs at the end points of the simulations for the various transmission pathways on humans, rodents and viruses

| Pathways using human force of infection |  |  |  |
| --- | --- | --- | --- |
| Pathways | Effect on Humans | Effect on Rodents | Effect on Virus |
| $I_R$ through $\beta_H$ | 8391 | $1.837 \times 10^4$ | $3.827 \times 10^6$ |
| $I_{HA}$ | 3007 | $1.462 \times 10^4$ | $1.425 \times 10^6$ |
| $V_S$ through $\eta_3$ | 943.7 | $1.299 \times 10^4$ | $6.556 \times 10^5$ |
| $I_{HS}$ | 22.55 | $1.185 \times 10^4$ | $2.445 \times 10^5$ |
| $V_A$ | 164.4 | $1.206 \times 10^4$ | $3.139 \times 10^5$ |
| $I_R$ & $I_{HS}$ | 8720 | $1.856 \times 10^4$ | $3.977 \times 10^6$ |
| $I_R$ & $I_{HA}$ | $1.518 \times 10^4$ | $2.158 \times 10^4$ | $6.828 \times 10^6$ |
| $I_R$ & $V_S$ | $1.253 \times 10^4$ | $2.042 \times 10^4$ | $5.634 \times 10^6$ |
| $I_R$ & $V_A$ | 9785 | $1.909 \times 10^4$ | $4.424 \times 10^6$ |
| $I_{HS}$ & $I_{HA}$ | 3611 | $1.504 \times 10^4$ | $1.652 \times 10^6$ |
| $I_{HS}$ & $V_S$ | 1131 | $1.319 \times 10^4$ | $7.356 \times 10^5$ |
| $I_{HS}$ & $V_A$ | 208.1 | $1.213 \times 10^4$ | $3.364 \times 10^5$ |
| $I_{HA}$ & $V_S$ | $1.126 \times 10^4$ | $1.905 \times 10^4$ | $4.652 \times 10^6$ |
| $I_{HA}$ & $V_A$ | 5621 | $1.617 \times 10^4$ | $2.371 \times 10^6$ |
| $V_S$ & $V_A$ | 2032 | $1.395 \times 10^4$ | $1.079 \times 10^6$ |
| $I_R$ & $I_{HS}$ & $I_{HA}$ | $1.554 \times 10^4$ | $2.177 \times 10^4$ | $7.013 \times 10^6$ |
| $I_R$ & $I_{HS}$ & $V_S$ | $1.292 \times 10^4$ | $2.062 \times 10^4$ | $5.825 \times 10^6$ |
| $I_R$ & $I_{HS}$ & $V_A$ | $1.015 \times 10^4$ | $1.929 \times 10^4$ | $4.595 \times 10^6$ |
| $I_R$ & $I_{HA}$ & $V_S$ | $1.82 \times 10^4$ | $2.294 \times 10^4$ | $8.32 \times 10^6$ |
| $I_R$ & $I_{HA}$ & $V_A$ | $1.639 \times 10^4$ | $2.213 \times 10^4$ | $7.416 \times 10^6$ |
| $I_R$ & $V_S$ & $V_A$ | $1.398 \times 10^4$ | $2.109 \times 10^4$ | $6.298 \times 10^6$ |
| $I_{HS}$ & $I_{HA}$ & $V_S$ | $1.208 \times 10^4$ | $1.951 \times 10^4$ | $5.048 \times 10^6$ |
| $I_{HS}$ & $I_{HA}$ & $V_A$ | 6527 | $1.668 \times 10^4$ | $2.721 \times 10^6$ |
| $I_{HS}$ & $V_S$ & $V_A$ | 2437 | $1.428 \times 10^4$ | $1.238 \times 10^6$ |
| $I_{HA}$ & $V_S$ & $V_A$ | $1.362 \times 10^4$ | $2.027 \times 10^4$ | $5.778 \times 10^6$ |
| $I_R$ & $I_{HS}$ & $I_{HA}$ & $V_S$ | $1.845 \times 10^4$ | $2.307 \times 10^4$ | $8.457 \times 10^6$ |
| $I_R$ & $I_{HS}$ & $I_{HA}$ & $V_A$ | $1.671 \times 10^4$ | $2.229 \times 10^4$ | $7.585 \times 10^6$ |
| $I_R$ & $I_{HS}$ & $V_S$ & $V_A$ | $1.435 \times 10^4$ | $2.128 \times 10^4$ | $6.485 \times 10^6$ |
| $I_R$ & $I_{HA}$ & $V_S$ & $V_A$ | $1.9 \times 10^4$ | $2.329 \times 10^4$ | $8.724 \times 10^6$ |
| $I_{HS}$ & $I_{HA}$ & $V_S$ & $V_A$ | $1.425 \times 10^4$ | $2.064 \times 10^4$ | $6.117 \times 10^6$ |
| $I_R$ & $I_{HS}$ & $I_{HA}$ & $V_S$ & $V_A$ | $1.922 \times 10^4$ | $2.34 \times 10^4$ | $8.847 \times 10^6$ |
| Pathways using rodent force of infection |  |  |  |
| Pathways | Effect on Humans | Effect on Rodents | Effect on Virus |
| $I_R$ through $\beta_R$ | $1.785 \times 10^4$ | $1.106 \times 10^4$ | $8.151 \times 10^6$ |
| $V_S$ through $\xi_1$ | $1.543 \times 10^4$ | 3233 | $6.738 \times 10^6$ |
| $I_R$ & $V_S$ | $1.922 \times 10^4$ | $2.34 \times 10^4$ | $8.847 \times 10^6$ |

## No Image files

## References

ACDC (2018). Lassa fever. Report on Lassa Fever: Centre for Disease Control and Prevention. Retrieved on 21-04-2021 from https://africacdc.org/disease/lassa-fever/.

Akhmetzhanov, A. R., Y. Asai, and H. Nishiura (2019). Quantifying the seasonal drivers of transmission for lassa fever in nigeria. Philosophical Transactions of the Royal Society B 374 (1775), 20180268.

Bausch, D. G., C. M. Hadi, S. H. Khan, and J. J. Lertora (2010). Review of the literature and proposed guidelines for the use of oral ribavirin as postexposure prophylaxis for lassa fever. Clinical infectious diseases 51 (12), 1435–1441.

Blower, S. M. and H. Dowlatabadi (1994). Sensitivity and uncertainty analysis of complex models of disease transmission: an hiv model, as an example. International Statistical Review/Revue Internationale de Statistique, 229–243.

Castillo-Chavez, C., Z. Feng, W. Huang, et al. (2002). On the computation of ro and its role on. Mathematical approaches for emerging and reemerging infectious diseases: an introduction 1, 229.

CDC (2014). Lassa fever transmission. Report on Viral Hemorrhagic fevers: US Department of Health and Human Services. Accessed on 21-04-2021 from https://www.cdc.gov/vhf/lassa/transmission/index.html.

Chitnis, N., J. M. Hyman, and J. M. Cushing (2008). Determining important parameters in the spread of malaria through the sensitivity analysis of a mathematical model. Bulletin of mathematical biology 70 (5), 1272–1296.

Control, P. P. (2018). How fast do mice multiply in your home? Preventive Pest Control. Retrieved on 21-02-2022 from https://www.preventivepesthouston.com/blog/2018/may/how-fast-do-mice-multiply-in-your-home-/.

Davies, J., K. Lokuge, and K. Glass (2019). Routine and pulse vaccination for lassa virus could reduce high levels of endemic disease: A mathematical modelling study. Vaccine 37 (26), 3451–3456.

Fichet-Calvet, E., B. Becker-Ziaja, L. Koivogui, and S. Günther (2014). Lassa serology in natural populations of rodents and horizontal transmission. Vector-Borne and Zoonotic Diseases 14 (9), 665–674.

Gibb, R., L. M. Moses, D. W. Redding, and K. E. Jones (2017). Understanding the cryptic nature of lassa fever in west africa. Pathogens and global health 111 (6), 276–288.

Gonzalez, S. G. (2020). Lassa fever. MedicineNet. Retrieved on 21-04-2021 from https://www.medicinenet.com/lassafever/article.htm.

Ibrahim, M. A. and A. Dénes (2021). A mathematical model for lassa fever transmission dynamics in a seasonal environment with a view to the 2017–20 epidemic in nigeria. Nonlinear Analysis: Real World Applications 60, 103310.

Lakshmikantham, V., S. Leela, and A. A. Martynyuk (1989). Stability analysis of nonlinear systems. Springer.

Ledder, G. (2017). Scaling for dynamical systems in biology. Bulletin of mathematical biology 79 (11), 2747–2772.

Lehmann, C., M. Kochanek, D. Abdulla, S. Becker, B. Böll, A. Bunte, D. Cadar, A. Dormann, M. Eickmann, P. Emmerich, et al. (2017). Control measures following a case of imported lassa fever from togo, north rhine westphalia, germany, 2016. Eurosurveillance 22 (39), 17–00088.

Lo Iacono, G., A. A. Cunningham, E. Fichet-Calvet, R. F. Garry, D. S. Grant, S. H. Khan, M. Leach, L. M. Moses, J. S. Schieffelin, J. G. Shaffer, et al. (2015). Using modelling to disentangle the relative contributions of zoonotic and anthroponotic transmission: the case of lassa fever. PLoS neglected tropical diseases 9 (1), e3398.

Marino, S., I. B. Hogue, C. J. Ray, and D. E. Kirschner (2008). A methodology for performing global uncertainty and sensitivity analysis in systems biology. Journal of theoretical biology 254 (1), 178–196.

Mikucki, M. A. (2012). Sensitivity analysis of the basic reproduction number and other quantities for infectious disease models. Ph. D. thesis, Colorado State University.

Moghadas, S. M., A. B. Gumel, R. G. McLeod, and R. Gordon (2003). Could condoms stop the aids epidemic? Journal of theoretical medicine 5 (3-4), 171–181.

NCDC (2021). An update of lassa fever outbreak in nigeria. Weekly Report on Lassa fever outbreak in Nigeria. Retrieved on 19-11-2021 from https://www.ncdc.gov.ng/diseases/sitreps/.

NCDC (2022). An update of lassa fever outbreak in nigeria. Weekly Report on Lassa fever outbreak in Nigeria. Retrieved on 15-03-2022 from https://www.ncdc.gov.ng/diseases/sitreps/.

Newman, T. (2018). Everything you need to know about lassa fever. MedicalNewsToday. Accessed on 21-04-2021 from https://www.medicalnewstoday.com/articles/306886.

NICD (2020). Lassa fever disease. Lassa Fever Report: Division of the National Health Laboratory Service Retrieved on 21-04-2021 from https://www.nicd.ac.za/diseases-a-z-index/lassa-fever/.

Obabiyi, O. and A. A. Onifade (2017). Mathematical model for lassa fever transmission dynamics with variable human and reservoir population. International Journal of Differential Equations and Applications 16 (1).

Ojo, M. M., B. Gbadamosi, T. O. Benson, O. Adebimpe, and A. Georgina (2021). Modeling the dynamics of lassa fever in nigeria. Journal of the Egyptian Mathematical Society 29 (1), 1–19.

Onah, I. S., O. C. Collins, P.-G. U. Madueme, and G. C. E. Mbah (2020). Dynamical system analysis and optimal control measures of lassa fever disease model. International Journal of Mathematics and Mathematical Sciences 2020.

Onuorah, M., M. Ojo, D. Usman, and A. Ademu (2016). Basic reproductive number for the spread and control of lassa fever. International Journal of Mathematics Trends and Technology (IJMTT) 30 (1), 1–7.

Ossai, E. N., O. E. Onwe, N. P. Okeagu, A. L. Ugwuoru, T. K. Eze, and A. S. Nwede (2020). Knowledge and preventive practices against lassa fever among heads of households in abakaliki metropolis, southeast nigeria: A cross-sectional study. Proceedings of Singapore Healthcare 29 (2), 73–80.

Peter, O. J., A. I. Abioye, F. A. Oguntolu, T. A. Owolabi, M. O. Ajisope, A. G. Zakari, and T. G. Shaba (2020). Modelling and optimal control analysis of lassa fever disease. Informatics in Medicine Unlocked 20, 100419.

Punch (2022). Lassa fever kills four doctors, infects 38 health workers. PUNCH Newspaper. Retrieved on 15-03-2022 from https://punchng.com/lassa-fever-kills-four-doctors-infects-38-health-workers/.

Richmond, J. K. and D. J. Baglole (2003). Lassa fever: Epidemiology, clinical features, and social consequences. Bmj 327 (7426), 1271–1275.

Sabeti, P. C. (2015). Clinical sequencing uncovers origins and evolution of lassa virus. Cell.

Saltelli, A., M. Ratto, T. Andres, F. Campolongo, J. Cariboni, D. Gatelli, M. Saisana, and S. Tarantola (2008). Global sensitivity analysis: the primer. John Wiley & Sons.

Saltelli, A., S. Tarantola, F. Campolongo, M. Ratto, et al. (2004). Sensitivity analysis in practice: a guide to assessing scientific models. Chichester, England.

Stephenson, E. H., E. W. Larson, and J. W. Dominik (1984). Effect of environmental factors on aerosol-induced lassa virus infection. Journal of medical virology 14 (4), 295–303.

Tewogbola, P. and N. Aung. Lassa fever: History, causes, effects, and reduction strategies. virus 2, 16.

Trends, M. (2021). Nigeria life expectancy 1951-2021. Macro Trends. Retrieved on 19-11-2021 from https://www.macrotrends.net/countries/NGA/nigeria/life-expectancy.

Turányi, T. (1990). Sensitivity analysis of complex kinetic systems. tools and applications. Journal of mathematical chemistry 5 (3), 203–248.

Van den Driessche, P. and J. Watmough (2002). Reproduction numbers and sub-threshold endemic equilibria for compartmental models of disease transmission. Mathematical biosciences 180 (1-2), 29–48.

Velasco-Hernandez, J. X. and Y.-H. Hsieh (1994). Modelling the effect of treatment and behavioral change in hiv transmission dynamics. Journal of mathematical biology 32 (3), 233–249.

WHO (2017). Lassa fever. WHO Factsheets. Accessed on 21-04-2021 from http://www.who.int/mediacentre/factsheets/fs179/en/.

WHO (2020). Lassa fever-nigeria. WHO Factsheets. Retrieved on 21-04-2021 from https://www.who.int/csr/don/20-february-2020-lassa-fever-nigeria/en/.

WHO (2021). Stakeholders shore up sensitization campaign to curb lassa fever outbreak in edo state. World Health Organization. Retrieved on 19-11-2021 from https://www.afro.who.int/news/stakeholders-shore-sensitization-campaign-curb-lassa-fever-outbreak-edo-state.

Yun, N. E. and D. H. Walker (2012). Pathogenesis of lassa fever. Viruses 4 (10), 2031–2048.

